# Cross study transfer of vascular transcriptomic classifiers in seven public cohorts

**DOI:** 10.64898/2026.10.29.26366574

**Authors:** Lingsen You, Yujun Guo, Xinyu Zhong, Zisu Peng, Wentong Wang, Li Shen, Junbo Ge

## Abstract

Strong discrimination within a vascular transcriptomic study may not transfer across clinical endpoints, tissue compartments or assay processing. We evaluated seven public Gene Expression Omnibus datasets containing 294 deposited observations, including repeated lesion segments. Four conventional classifiers and a prior-only control underwent nested validation in 48 abdominal aortic aneurysm samples, followed by external evaluation under three preprocessing configurations. Source-rank L1 logistic regression achieved an area under the receiver operating characteristic curve of 0.947; external values were 0.753 and 0.667 in two carotid cohorts and 1.000 in 11 macrophage plaque samples. The latter value fell to 0.367 under a separately source-tuned common-gene pipeline. In 126 plaque donors with censored follow-up, Harrell concordance was 0.565 (95% interval 0.444–0.678) and 0.524 after a 30-day landmark. Processing-incompatible blood data served as a stress control; 43 correlated plaque segments were analysed descriptively. A matched permutation test supported source separation without testing generalisation. These results show that internal discrimination and isolated external success do not establish stable panvascular risk prediction. The benchmark supplies code, per-sample outputs and provenance for scrutinising pipeline sensitivity, endpoint mismatch and observation dependence.

## Introduction

A transcriptomic classifier can distinguish samples within one vascular study while failing in another. Different vascular beds share inflammatory and remodelling processes, but their tissue composition, sampling procedures, clinical phenotypes and expression assays differ. For a model intended to support panvascular research, the relevant question is whether discrimination persists when these conditions change. A strong internal area under the receiver operating characteristic curve (AUROC) is insufficient evidence of that transfer.

Cross-study validation evaluates prediction under study-level distribution shifts that random within-study partitions do not reproduce [1]. In high-dimensional expression data, selecting genes before cross-validation can also create optimistic estimates [2]. Model tuning must be separated from performance estimation [3], and uncertainty is particularly important when the numbers of cases and controls are small [4]. Independent investigations of transcriptomic preprocessing have further shown that choices which appear reasonable within one dataset can behave differently across studies [5]. These issues motivate a benchmark that records both the statistical fitting boundary and the biological meaning of each external endpoint.

Vascular intervention research adds a specific interpretation problem. A proposed tri-ecological framework for vascular-device matching highlights the interaction of mechanical, cellular and physicochemical-immune conditions across vascular beds [6]. This provides a conceptual motivation to examine context dependence; it does not establish the transferability of a transcriptomic classifier. Ruptured abdominal aortic aneurysm, symptomatic carotid stenosis, intraplaque haemorrhage, future ischaemic events and peripheral artery disease are related clinical contexts, but they are not interchangeable outcomes. An intervention-outcome study, such as the reported intra-aortic balloon pump analysis [7], provides clinical motivation for careful evaluation; it cannot validate a transcriptomic classifier or establish that the classifier identifies treatment benefit. We therefore treat each public vascular dataset as a separately specified transfer question.

We developed VascBridge-Instability as an executable benchmark of source discrimination and external transfer. This revision expands an earlier four-cohort exploratory analysis with three public datasets, compares conventional learning algorithms under nested source-cohort validation, and audits gene coverage, preprocessing, aneurysm-size structure and the unit of observation. Its contribution is a documented computational evaluation procedure and its empirical failure analysis. It does not introduce a foundation model or a clinically deployable risk score.

## Methods

### Design and public data selection

Only openly accessible Gene Expression Omnibus (GEO) expression files, platform annotations, sample metadata and original publications were used [8, 9]. The development cohort was fixed as GSE98278. The original external studies were GSE111782, GSE198600 and GSE27034; their earlier results had already been examined. Analyses on these three studies are therefore explicitly revision analyses, not pristine prospective or untouched validation. Additional studies were selected by public metadata and biological relevance before the revised score results were examined. No external labels were used for hyperparameter tuning, and every model variant in the dated revision configuration is reported.

Cohort selection was purposive, not an exhaustive systematic review. Eligible records had publicly downloadable processed human vascular expression, a recoverable clinical or pathological label, and an interpretable relationship to the source question. GSE41571 supplied ruptured and stable carotid plaque macrophages. GSE21545 supplied carotid plaque expression with follow-up event status and time. GSE163154 supplied intraplaque haemorrhage (IPH) and non-IPH segments. The candidate GSE100927 was not added because atherosclerotic-versus-donor arterial tissue addresses a different question and would not remedy the lack of an independent, identically defined aneurysm rupture cohort.

Each input file is linked to its public accession, retrieval record and SHA256 digest. Expression columns are checked against sample identifiers before label assignment. The audit distinguishes samples, lesion segments and identified donors, excludes paired blood arrays from the plaque survival dataset, and records study-specific exclusions. No shared GEO sample identifiers were found across the included studies. Different recruiting institutions and publications reduce the likelihood of duplicated cohorts, but anonymised public records cannot rule out all cross-study participant overlap.

The cohort records are individually identified as GSE98278 [10], GSE111782 [11], GSE198600 [12], GSE27034 [13], GSE41571 [14], GSE21545 [15] and GSE163154 [16]. The analysis configuration, amendments and source audit are provided in Supplementary Data 1.

Related work by the authors evaluates matched comparators for longitudinal vascular transcriptome module prediction (VascFlux-GEO) and anatomical comparison populations and threshold transport (VascTransport). The present study instead evaluates classifiers trained on aneurysm rupture status across distinct vascular endpoints. GSE27034 is shared with the descriptive component of VascFlux-GEO but is used here only as a processing-incompatible stress control; those module contrasts are not presented as new findings in this manuscript.

### Ethics approval and consent to participate

This secondary analysis used openly available, de-identified public expression and repository metadata. It involved no new recruitment, specimen collection or attempt to re-identify participants. No additional ethics approval or participant consent was obtained for this public-data reanalysis. The original source studies describe their recruitment and ethical oversight; their approvals are not represented as new approvals for this computational study.

### Endpoints and units of observation

GSE98278 contributes 48 aneurysm tissue samples, comprising 31 elective stable and 17 ruptured specimens. The outcome represents status at tissue sampling, rather than subsequent rupture. GSE111782 includes nine symptomatic and nine asymptomatic carotid plaques. GSE198600 includes six plaques associated with a recent carotid-related ischaemic event and five asymptomatic plaques. GSE27034 comprises 19 peripheral artery disease and 18 control peripheral blood mononuclear-cell samples; it is a systemic disease-status stress test rather than lesion-rupture validation.

GSE41571 contains five ruptured and six stable carotid plaques analysed using laser-microdissected macrophages. This provides a rupture-related endpoint while changing both vascular bed and sampled cellular compartment. GSE21545 contains 126 plaque samples with unique deposited patient identifiers, 25 subsequent ischaemic events and 101 censored observations. Its 97 paired blood samples were excluded. Event status was not relabelled as baseline symptomatic disease. Survival discrimination was analysed using the recorded follow-up times; a 30-day landmark analysis excludes the 12 events within 30 days and uses 114 remaining donors with 13 events.

The original GSE163154 publication describes paired lesion segments from 24 symptomatic patients, whereas GEO deposits 43 transcriptomic samples (27 IPH and 16 non-IPH). The original multi-omics analysis used 42 segments after one sample failed mass-spectrometry criteria; that exclusion is not evidence of failed expression measurement. A donor-to-GEO-sample crosswalk was not available. We retained the 43 deposited expression samples for descriptive segment-level scoring, did not treat them as 43 independent patients, and did not calculate patient-level confidence intervals or nominal significance tests for this cohort.

The original reports support the macrophage sampling procedure [17], the censored post-endarterectomy outcome [18], and the repeated-segment structure of the IPH dataset [19]. Table 1 summarises the cohorts and their evaluation roles.

**Table 1.** Cohorts and endpoint-specific evaluation roles.

| Accession | Material and endpoint | Included observations | Interpretation |
| --- | --- | --- | --- |
| GSE98278 | AAA tissue; ruptured / elective stable | 48; 17 / 31 | Source development |
| GSE111782 | Carotid plaque; symptomatic / asymptomatic | 18; 9 / 9 | Previously examined endpoint shift |
| GSE198600 | Carotid RNA-seq; recent event / asymptomatic | 11; 6 / 5 | Previously examined endpoint and assay shift |
| GSE41571 | Macrophages; ruptured / stable carotid plaque | 11; 5 / 6 | Rupture-related cell-compartment shift |
| GSE21545 | Carotid plaque; future ischaemic event / censored | 126 donors; 25 / 101 | Survival discrimination; not a rupture endpoint |
| GSE163154 | Carotid IPH / non-IPH lesion segments | 43 segments; 27 / 16 | 24 source patients; descriptive only |
| GSE27034 | PAD / control PBMCs | 37; 19 / 18 | Preprocessing-incompatible stress control |

### Probe mapping and processing compatibility

Microarray probes were mapped using the corresponding public GEO platform annotations. Probes with no symbol or more than one unique symbol were excluded, and the median combined probes mapping to the same symbol. This stricter rule was applied consistently across old and new microarray datasets. RNA-seq FPKM values were reconstructed from the public GSE198600 supplementary archive, collapsed by gene identifier, and checked against all eleven metadata sample identifiers. No pooled batch correction or external-label-based normalisation was applied.

Numerical values were inspected rather than assuming the metadata scale descriptions were sufficient. GSE98278 contained background-corrected values including negatives; its primary transformation used ranks and did not assume log2 intensity. The GSE41571 series matrix contained probe-median ratios. We instead used the eleven uncentred processed-intensity columns labelled raw in the official supplementary matrix, mapped them by deposited specimen/CEL identity and checked the companion normalised columns against all 601,425 corresponding series-matrix entries. These are processed intensities, not unprocessed CEL measurements. GSE27034 had an across-sample median of zero for all 54,675 probes, making within-sample ranks incompatible with abundance ranks in the source. PAD is therefore retained only as a processing stress control, outside biological transfer claims. The known impact of technical processing motivates this separation [20].

### Source-derived representation and nested model fitting

The primary feature vocabulary comprised 20,748 unambiguously annotated genes in the source cohort alone. For each sample, observed genes within that vocabulary were ranked and mapped to (rank - 1)/(number observed - 1). Ties received average ranks. Absent genes remained missing and were replaced with medians learned from the source-training partition; they were never represented as measured zero expression. Rank denominators consequently depend on measured gene coverage. This design preserves source-only feature eligibility but does not guarantee cross-platform measurement equivalence.

Inside every inner training partition, a Pipeline fitted median imputation, removal of constant features, a maximum 5,000-gene variance shortlist, ANOVA selection of 25 or 100 genes, standard scaling and the classifier. L1 logistic regression [21], elastic-net logistic regression (L1 ratio 0.5), linear support vector machine (SVM), random forest and a source-prevalence-only dummy model were compared. Linear-model regularisation C was selected from 0.1 and 1.0. The forest used 200 trees, a minimum leaf size of two, square-root feature subsampling and a maximum depth of three or unrestricted depth. Grid selection used three-fold source-only AUROC; ties followed the fixed grid order. The executable configuration specifies solvers, stopping limits and seeds [22].

The outer validation used stratified four-fold cross-validation repeated three times, with each source sample appearing in a test fold once per repeat. No outer test row entered inner fitting. The primary internal statistic used one score per sample obtained by averaging its three out-of-fold (OOF) predictions; 144 predictions were not counted as 144 independent observations. Each algorithm was then tuned on the full source through three-fold validation and refitted on all source samples for external scoring. No external score threshold or probability recalibration was fitted. This explicitly separates hyperparameter selection from internal performance estimation [3, 23].

### Pipeline and size sensitivity analyses

Two separately source-tuned L1 pipelines supplemented the primary analysis. The common-rank pipeline used 11,845 genes measured in the source and all three previously examined external studies. This vocabulary uses external feature availability without external labels and is explicitly transductive. It changes rank denominators, candidate genes, selected features and the refitted model jointly. It is a pipeline sensitivity analysis, not an isolated causal test of vocabulary or missingness. A scale diagnostic used signed log2(1 + absolute intensity) for source values, log2(1 + nonnegative intensity) for GSE41571 and GSE198600 FPKM, and retained the other deposited processed scales. Source-fitted standardisation was used. Because those scales are not necessarily comparable, this is a diagnostic rather than a validated alternative cross-platform normalisation.

Public aneurysm-size categories contained 15 intermediate samples (all stable), 16 large samples (seven stable, nine ruptured) and 17 with unknown size (nine stable, eight ruptured). We evaluated a fixed regularised logistic model using only the category indicators under the same outer folds. Two different subgroup questions were reported: nested retraining restricted to the large or unknown-size subset, and evaluation within those categories of OOF scores learned using the full-source training folds. The latter was added as a reporting audit to avoid interpreting the restricted-retraining contrast as a pure diameter effect. The intermediate category could not support binary subgroup discrimination because it contained no ruptures.

### Metrics and uncertainty

Binary endpoints were reported separately with AUROC and average precision (the stepwise precision-recall summary used here) [24]. Brier scores were calculated only for models producing probabilities, and are descriptive because source-event probabilities are not calibrated to a different endpoint [25]. SVM margins were not converted into probabilities for Brier scoring. Two thousand class-stratified bootstrap resamples of saved scores provided percentile AUROC intervals [26]. These intervals are conditional on fitted/OOF scores, not full model-development uncertainty; source resampling also assumes sample independence that could not be verified from donor identifiers. Degenerate intervals from perfect separation or constant scores are flagged rather than reported as informative confidence bounds. IPH segment-level estimates have no patient-level interval.

For GSE21545, higher frozen classifier scores were interpreted as higher hypothesised event risk solely to test directional survival discrimination. Harrell concordance uses pairs where the earlier observation has an observed event, assigning half credit to tied scores [27]. Event and censoring times were retained; no censored individual was treated as a known permanently event-free control. Two thousand donor-bootstrap resamples produced conditional intervals. The 30-day landmark, specified during endpoint auditing before score inspection, excluded observations ending at or before day 30 and reset the remaining follow-up origin. Harrell concordance depends on the censoring distribution, and no proportional-hazards or treatment-effect inference was attempted.

A separate fixed L1 baseline (100 selected genes, C=0.25) underwent 200 label permutations. The complete train-only preprocessing and fitting pipeline was refitted for each fold, using the same four-fold, three-repeat stratified splitter rule for observed and permuted labels. The one-sided empirical P value was (1 + number of permuted AUROCs at least as large as observed)/(1 + 200) [28]. This tests that fixed baseline under an exchangeability assumption, not the separately tuned models, transportability, causal effects or absence of size confounding. No multiple-endpoint hypothesis-testing programme or external success threshold was used to select a model.

### Revision boundaries and reproducibility

The revision changes both preprocessing and validation relative to the earlier manuscript: ambiguous probe mappings are excluded, the primary vocabulary is source-derived, missing external genes are imputed with source-training medians, and hyperparameters are selected within nested validation. Revised values therefore replace the earlier headline estimates; changes cannot be attributed solely to adding datasets. The dated configuration describes a revision analysis plan, not public preregistration. External cohorts are never pooled with the source for supervised training.

The baseline comparison is deliberately limited to conventional algorithms appropriate for a 48-sample development set. It is not a claim to outperform contemporary foundation models or all state-of-the-art methods. No winning method is selected from external AUROC. All source-selected models, including unsuccessful transfer and incomplete gene coverage, are included in the result tables.

Python 3.11.9, NumPy 2.4.6, pandas 2.3.3, SciPy 1.17.1 and scikit-learn 1.9.0 were used; dependencies, seeds, source hashes and software commands are archived. Independent internal checks recomputed 61 metric rows, 108 outer-fold records, 42 frozen model-by-external-cohort predictions and 200 permutation statistics. Those checks establish consistency of the supplied computation and do not constitute independent clinical replication. The reporting audit follows the relevant data, optimisation, model and evaluation categories of DOME [23].

### Computational provenance

OpenAI Codex assisted with research-planning drafts, source retrieval, development of the analysis and plotting scripts, and manuscript drafting and assembly. The reported numerical values were computed by the archived Python programs. Internal verification recomputed metrics from saved predictions and checked sample identities, partition boundaries and figure values against the archived outputs. These computational checks do not constitute external replication or peer review.

## DECLARATION OF GENERATIVE AI AND AI-ASSISTED TECHNOLOGIES

Codex assisted with LaTeX assembly, prose editing, and consistency checks. The authors reviewed and revised all AI-assisted text and take responsibility for the submitted manuscript.

## Results

### Public data audit and analysis coverage

The seven datasets contributed 294 deposited observations to distinct analyses, including 43 correlated IPH and non-IPH plaque segments and 126 identified plaque donors; this is not a pooled 294-patient validation cohort. Fig 1 shows the development boundary and endpoint-specific sample counts. Removing 97 paired blood arrays from GSE21545 avoided double-counting its donors. Recovering uncentred GSE41571 intensities and identifying the incompatible PAD matrix changed the interpretation of the earlier manuscript before biological transfer was assessed. All revised metrics, folds, predictions and missingness records are supplied in Supplementary Data 1.

**Fig 1.**
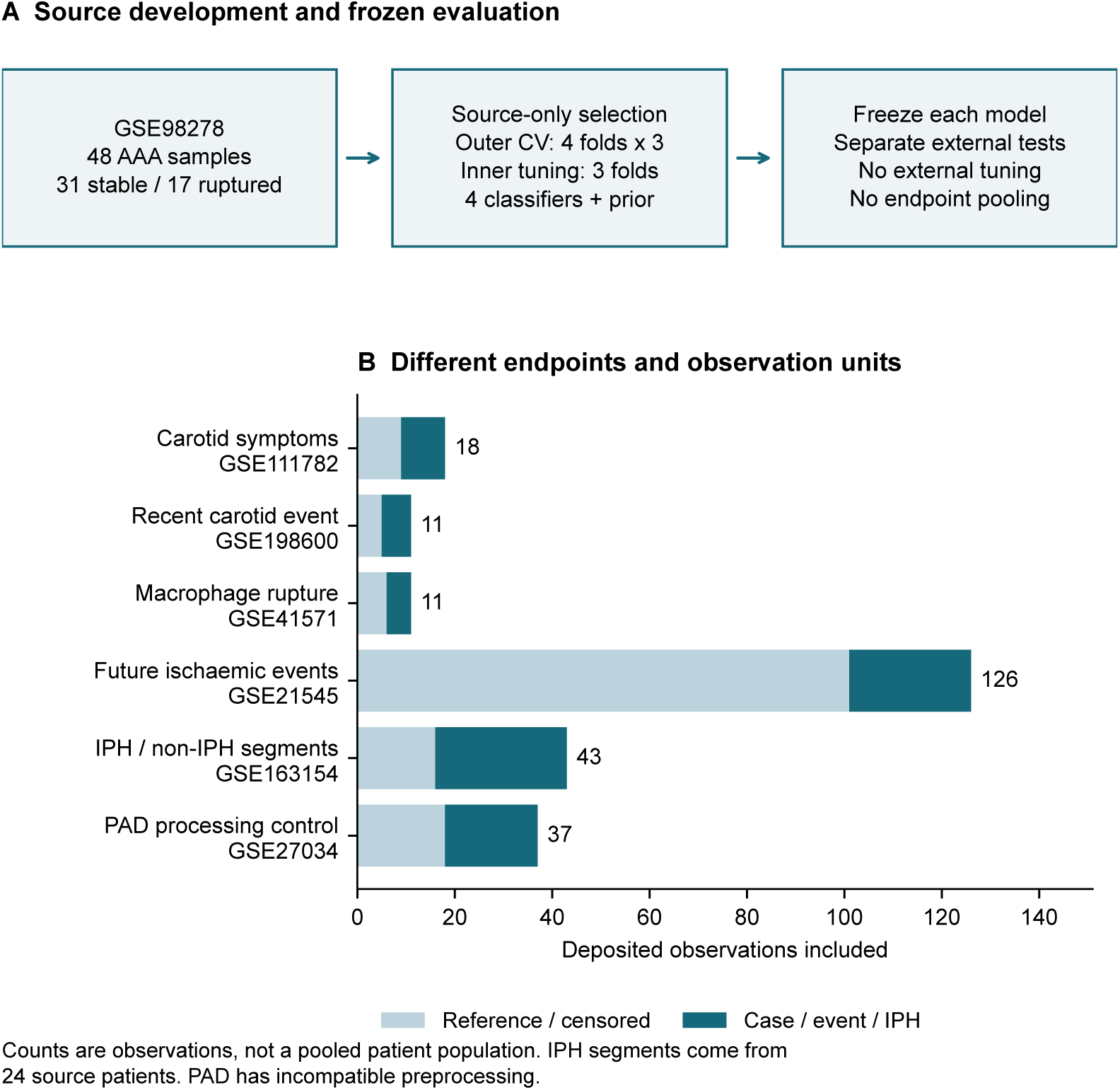
Study design and observation units. Source-only nested fitting was followed by separate external tests. Case labels represent distinct biological or follow-up endpoints; censored observations are not permanent negative controls. IPH segments are repeated observations from 24 source patients, and PAD is a processing-incompatible control. OpenAI Codex assisted with the plotting code; the figure was rendered by Matplotlib from the archived numerical outputs.

### Nested source discrimination and conventional baselines

Source-rank nested mean-OOF AUROCs were 0.947 for L1 logistic regression, 0.949 for elastic net, 0.918 for linear SVM and 0.911 for random forest (Supplementary Table S1 and Fig 2). L1 average precision was 0.915 and its conditional AUROC interval was 0.869-0.996. These results describe sample separation in the source cohort. The prior-only model had fold-level AUROC 0.500, but its averaged-OOF AUROC was 0.426 because small prevalence differences between training folds produced different constant scores. No learned discriminative information is implied by that numerical deviation from 0.500.

**Fig 2.**
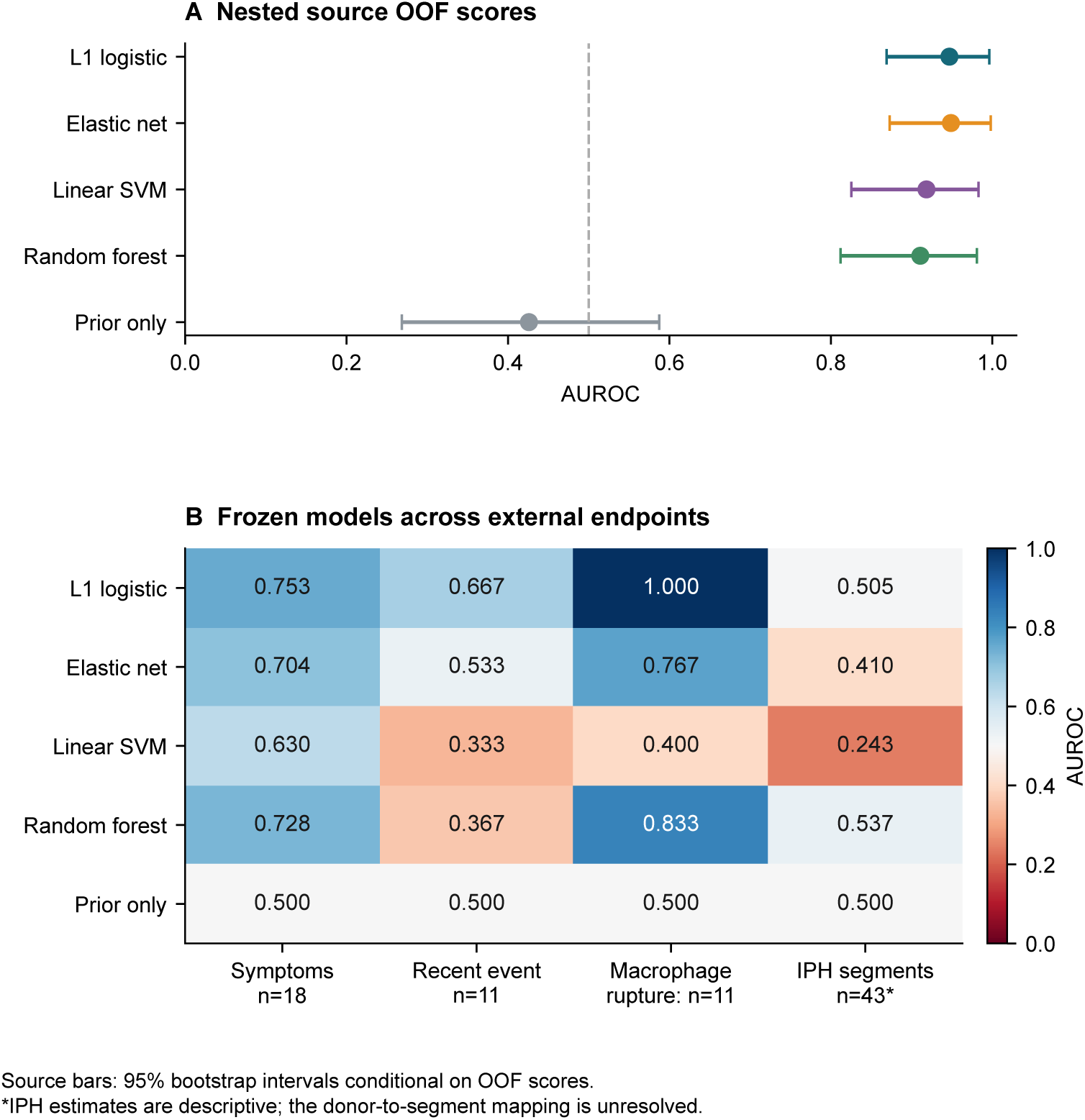
Nested internal and endpoint-specific external discrimination. Panel A shows conditional source-score bootstrap intervals. Panel B reports all conventional models, including failures; IPH results are descriptive segment-level scores. The 11-sample perfect ranking of one pipeline is not a population-level guarantee. OpenAI Codex assisted with the plotting code; the figure was rendered by Matplotlib from the archived numerical outputs.

### External binary endpoints and complete pipeline sensitivity

The source-tuned L1 model used C=0.1 and 25 ANOVA-selected genes, of which three coefficients were nonzero. Its external AUROCs were 0.753 (95% interval 0.494-0.963) for carotid symptoms, 0.667 (0.300-1.000) for recent-event RNA-seq, and 1.000 for the 11 macrophage samples (Table 2). IPH discrimination was 0.505, reported without an independence-based interval. Across the four trained primary algorithms, the macrophage-cohort AUROC ranged from 0.400 to 1.000. A single favourable model-dataset pair therefore did not represent model-independent replication.

**Table 2.** Main L1 scores on distinct external binary endpoints.

| Endpoint | n | AUROC | 95% interval | Average precision |
| --- | --- | --- | --- | --- |
| Carotid symptoms | 18 | 0.753 | 0.494-0.963 | 0.781 |
| Recent-event RNA-seq | 11 | 0.667 | 0.300-1.000 | 0.672 |
| Macrophage rupture | 11 | 1.000 | Not estimated* | 1.000 |
| IPH segments | 43 | 0.505 | Not estimated* | 0.700 |

\*The perfect-ranking macrophage bootstrap is degenerate and cannot supply an informative uncertainty interval; the IPH analysis lacks a donor crosswalk and therefore has no patient-level interval. These are different reasons for not reporting a numerical interval.

The common-rank pipeline produced a slightly higher source AUROC (0.956) but external AUROCs of 0.593, 0.400 and 0.367 in the same carotid array, RNA-seq and macrophage cohorts (Fig 3). Its source-selected settings were 100 genes and C=1.0, so this comparison changes vocabulary, ranking, selected features and fitted coefficients together. The scale diagnostic also gave a different pattern. These contrasts demonstrate sensitivity to complete modelling pipelines; they do not identify one causal source of the differences.

**Fig 3.**
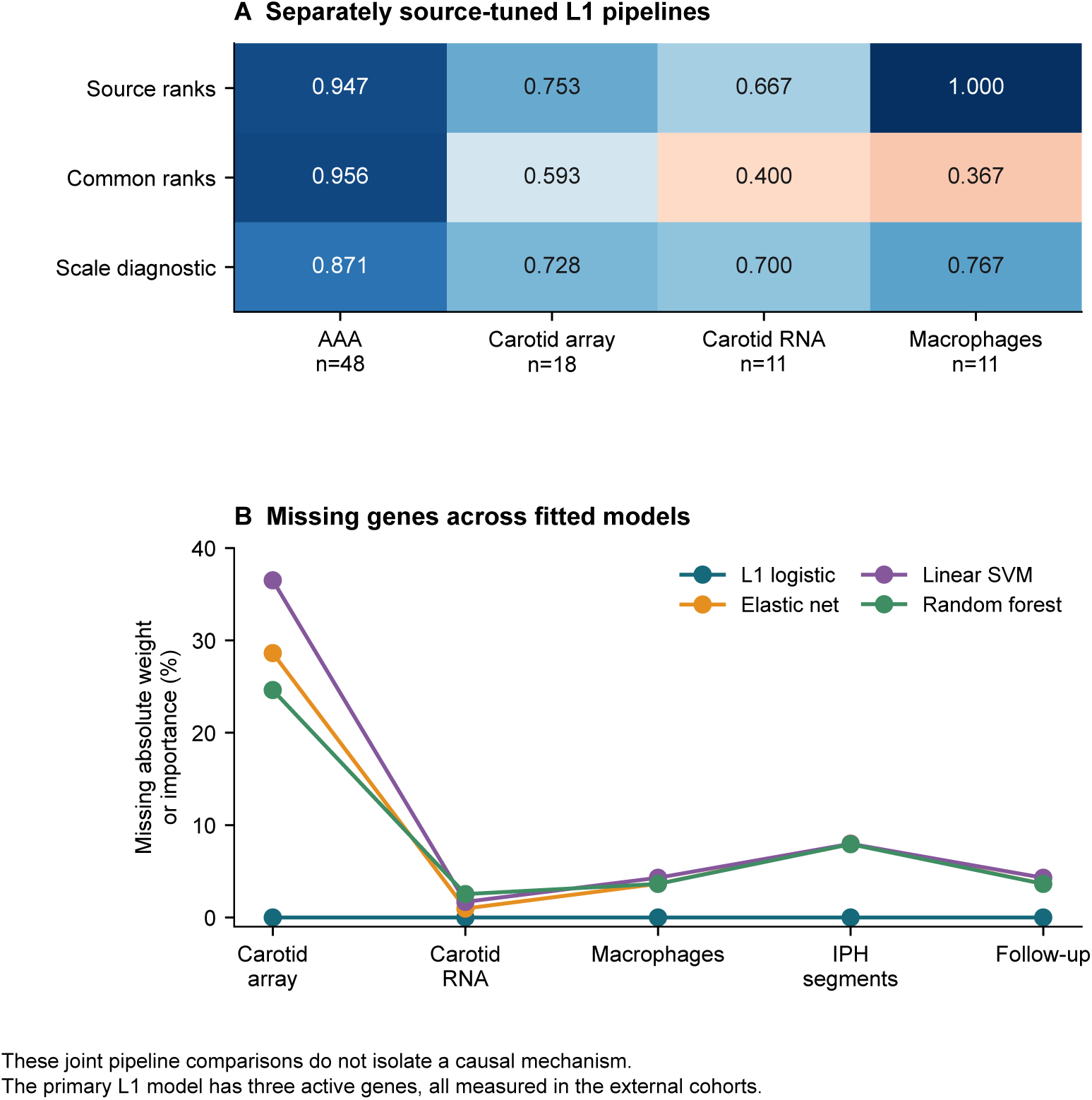
Complete pipeline sensitivity and active-feature coverage. Panel A compares separately tuned source-rank, common-rank and scale-diagnostic L1 pipelines. Panel B reports the fraction of absolute coefficient weight or forest feature importance associated with unavailable genes. Gene availability, rank denominators and refitting change jointly; this is not a single-factor ablation. OpenAI Codex assisted with the plotting code; the figure was rendered by Matplotlib from the archived numerical outputs.

Eight of the main L1 pipeline’s 25 ANOVA-selected features were absent from GSE111782, but all three nonzero-coefficient genes were measured in every external dataset. Thus missing active L1 features do not explain its transfer pattern. Other algorithms were more exposed to coverage differences: for the carotid array, absent genes represented 28.6% of elastic-net absolute coefficient weight. Coverage is a model-specific diagnostic, and rank denominators can change even when the active genes themselves are present. All individual selected and active-feature records are supplied.

### Censored follow-up discrimination

The main L1 score had Harrell concordance 0.565 (95% donor-bootstrap interval 0.444-0.678) across 126 plaque donors and 25 events. After excluding 12 events within 30 days, concordance was 0.524 (0.339-0.721) among 114 donors and 13 subsequent events. The corresponding full-follow-up values for elastic net, SVM and random forest were 0.565, 0.448 and 0.559 (Fig 4). Broad intervals and sensitivity to the observation period do not support a reliable prognostic claim. Full and landmark analyses share donors and are not independent replications.

**Fig 4.**
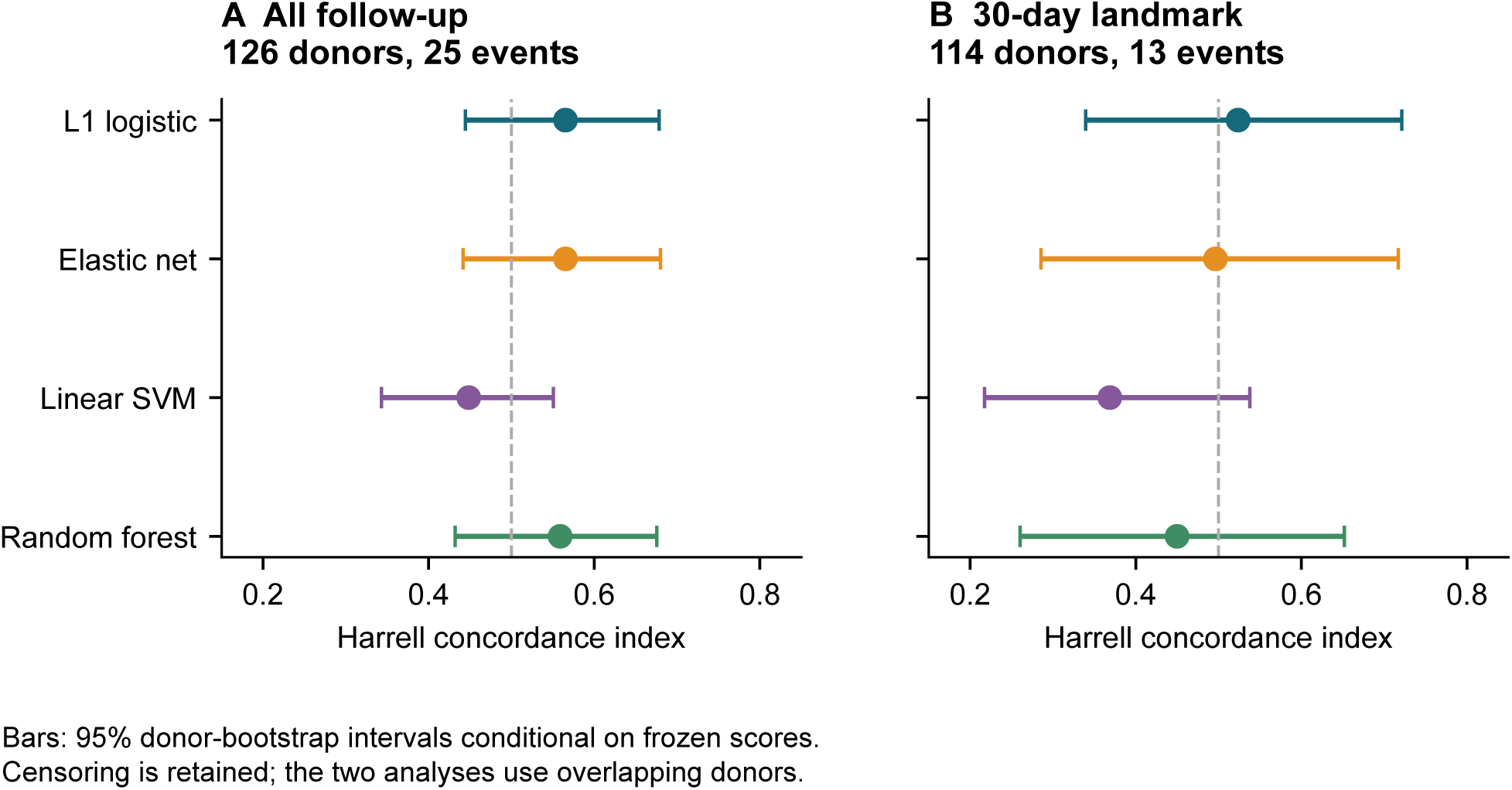
Survival discrimination in the plaque follow-up cohort. Frozen source-model scores were evaluated using event and censoring times. The 30-day landmark removes early events and resets the remaining follow-up origin. Error bars represent 2,000 donor-bootstrap resamples conditional on the saved scores; model-development uncertainty is not included. OpenAI Codex assisted with the plotting code; the figure was rendered by Matplotlib from the archived numerical outputs.

### Size structure and restricted retraining

The category-only source model achieved AUROC 0.673. Within the 16 large aneurysms, nested L1 retraining restricted to that subset achieved 0.246; the analogous value for the 17 unknown-size samples was 0.875. By contrast, evaluation of full-source mean-OOF scores within those same subsets gave 0.921 and 0.903, respectively (Fig 5). This contrast shows sensitivity to training size, composition and refitting. It does not show that adjustment for aneurysm diameter alone eliminates discrimination, nor does it establish absence of diameter confounding.

**Fig 5.**
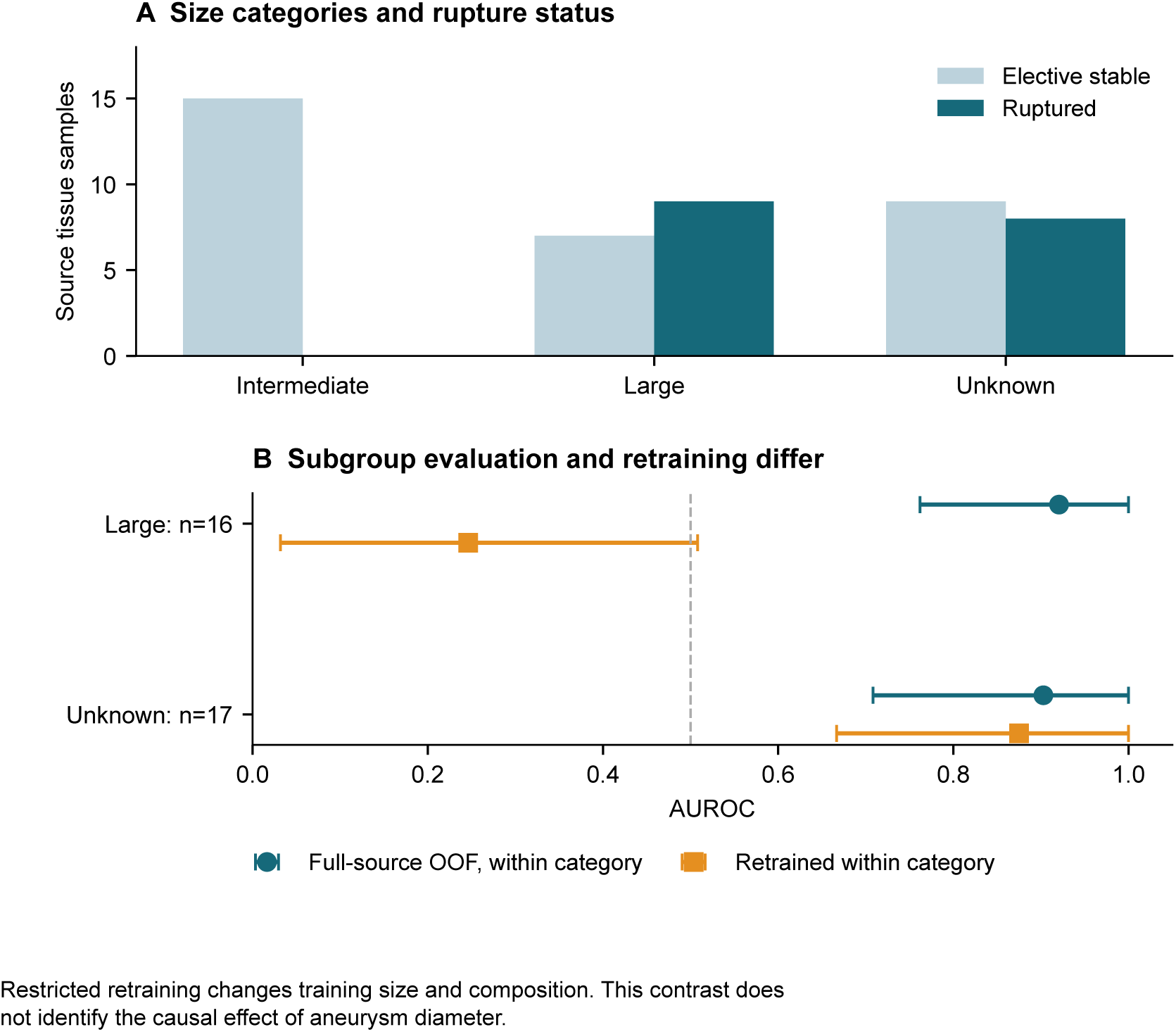
Size-category structure and two subgroup estimands. Panel A shows deposited label counts. Panel B separates evaluation of full-source OOF scores within a stratum from retraining a new model only within that stratum. The procedures differ in training-set size, composition and tuning; their contrast cannot identify a causal diameter effect. OpenAI Codex assisted with the plotting code; the figure was rendered by Matplotlib from the archived numerical outputs.

### Matched permutation control and limits of perfect ranking

The fixed, untuned permutation baseline yielded AUROC 0.941; the mean of 200 null AUROCs was 0.473, with an empirical 2.5th-97.5th percentile range of 0.239-0.708. No null value exceeded the observed statistic, giving empirical one-sided P=1/201=0.00498. This result concerns source separation by that fixed baseline only. In the macrophage cohort, the main tuned L1 pipeline perfectly ordered five cases and six controls, so all class-stratified bootstrap resamples also had AUROC 1.000. The resulting interval [1.000, 1.000] was non-informative about population uncertainty and was suppressed. Fig 6 displays the actual eleven scores rather than representing this small-sample result as certain or clinically calibrated.

**Fig 6.**
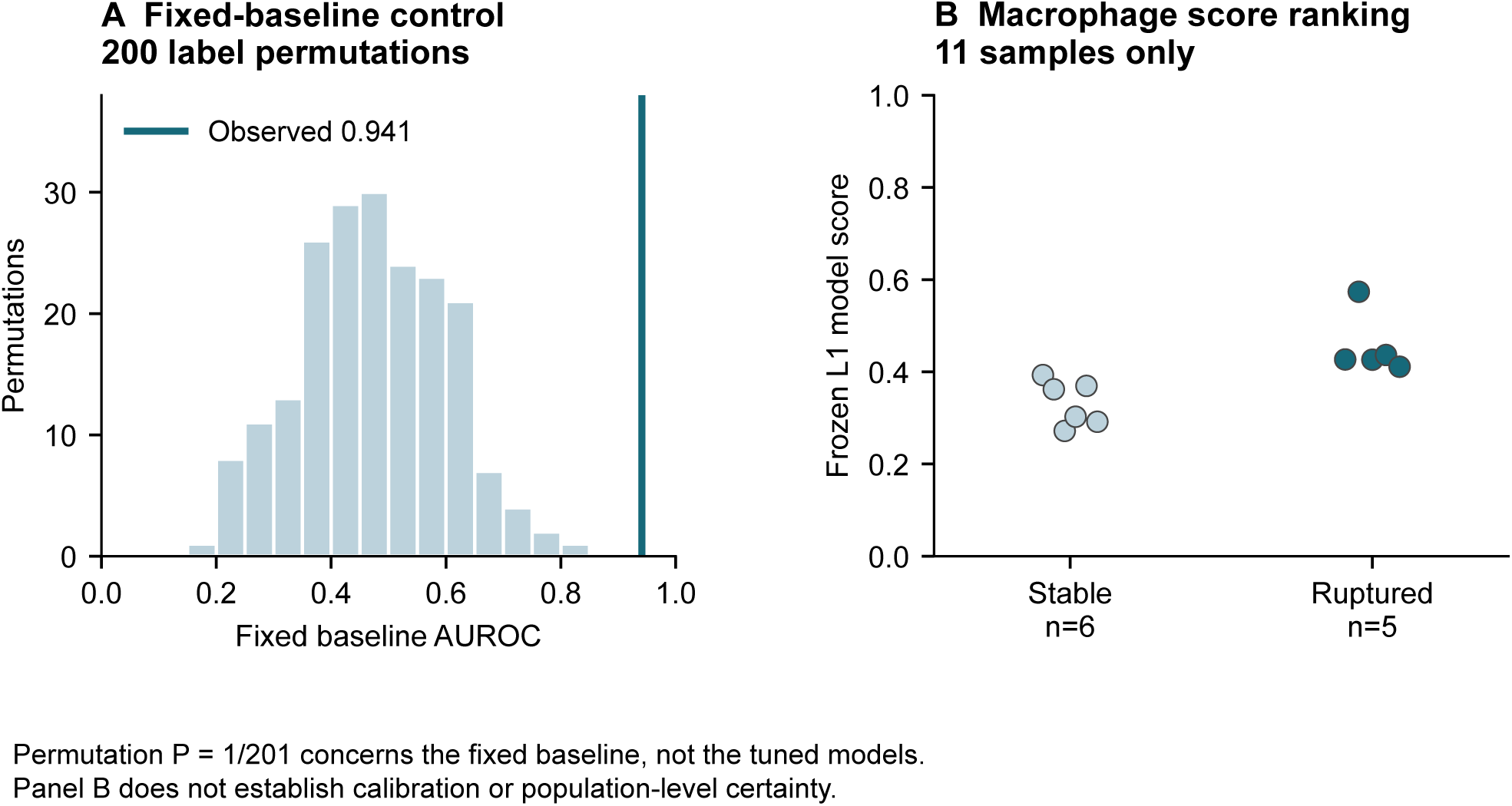
Fixed-baseline permutation control and small-cohort score distribution. Panel A reruns the complete selection and fitting procedure under 200 shuffled label vectors with matched validation rules. Panel B shows each of the eleven macrophage samples. Perfect ordering of these observed samples does not establish a validated threshold or transportable absolute risk. OpenAI Codex assisted with the plotting code; the figure was rendered by Matplotlib from the archived numerical outputs.

The PAD processing control is retained in the complete machine-readable results, with its incompatible-preprocessing role on every row. Its numerical AUROC is not counted as biological replication or failure. The original manuscript’s cross-endpoint success gate is not used in this revision, because the audited endpoints and observation units do not support a common success count.

## Discussion

This expanded public-data analysis found high source discrimination alongside external results that depended strongly on the full modelling pipeline and the endpoint being tested. The most striking result was not uniformly poor transfer: one source-only L1 pipeline perfectly ranked a small independent macrophage cohort, whereas other source-tuned models and the common-gene L1 pipeline did not. In the larger censored plaque cohort, discrimination was modest and uncertain. These findings favour reporting all specified model-dataset combinations over selecting the strongest external pair.

The important distinction is between reproducible computation and reproducible biological transfer. A deterministic pipeline can repeatedly produce a high internal AUROC without preserving the same relationship in another study. Reporting the external results alongside source performance and gene coverage makes this distinction inspectable. Weak transfer is informative about the tested models and datasets, but it does not prove that vascular beds lack shared mechanisms or that more suitable representations cannot transfer.

The dataset audit also changes what can legitimately be inferred. Counting the 43 GSE163154 segments as independent patients would manufacture precision. Treating the GSE21545 censored observations as uncomplicated negative controls would discard follow-up time. Pooling these endpoints into a single external success count would conflate biological questions. Separate endpoint and unit-of-observation records are therefore a substantive part of the benchmark rather than administrative metadata.

The comparison remains a conventional baseline study. Its purpose is to expose failure modes that future vascular AI methods should address under an identical evaluation protocol. Related work on intravascular imaging and image-derived digital twins also identifies data heterogeneity and limited multicentre validation as translational barriers [29]. That review provides a perspective on future multimodal applications, which were not evaluated in this transcriptomic benchmark. A useful next method would demonstrate improvements on biologically aligned independent cohorts, report feature coverage and calibration appropriate to the outcome, and preserve patient-level separation. This study provides no basis for selecting a vascular device, recommending an intervention, or predicting an individual patient’s treatment benefit.

The findings extend the practical concerns identified in cross-study prediction and expression-model validation [1, 5, 30]. They do not establish a new learning algorithm. The reusable contribution is the evaluation procedure: explicit endpoint roles, a sample-unit audit, processing compatibility checks, nested source tuning, complete external result reporting and conditionally interpreted uncertainty. For studies that later seek clinical prediction, additional outcome-specific development and reporting would be required; the present benchmark is not a completed clinical prediction model validation [31].

### Limitations

The source cohort is small, and aneurysm size is structurally associated with the rupture label. Its public size categories support only limited sensitivity analyses, not adjustment for continuous diameter, medication, cell composition or all clinical confounding. Even nested cross-validation cannot remove this confounding or substitute for an independent aneurysm rupture cohort. Existing cross-validation scores also share overlapping training sets, so sample bootstrap intervals are conditional descriptions and do not include the full uncertainty from model development.

External endpoints, tissues, assays and processing pipelines differ simultaneously. Differences in performance cannot be assigned causally to vascular bed, assay platform or biology. Public processed matrices retain the original studies’ preprocessing decisions; raw-data reprocessing and cross-platform biological calibration were not attempted. Probe annotations can be incomplete or outdated, and the strict mapping and source-median imputation rules are transparent conventions rather than proof of measurement equivalence.

The newly added rupture-related macrophage cohort remains small. Follow-up discrimination in the plaque cohort concerns subsequent composite ischaemic events, with substantial early postoperative events, rather than aneurysm rupture. The IPH cohort has unresolved within-patient mapping. Thus the expanded dataset collection increases the range of explicit tests but does not turn the benchmark into a large clinical validation study. No subgroup fairness assessment, clinical decision analysis, prospective deployment or experimental validation was performed.

The manuscript is a revision following examination of earlier external results. Although revised hyperparameters were selected only within the source cohort and the newly added endpoints were recorded before score inspection, analyst choices and the purposive selection of studies remain potential sources of bias. The code and complete result tables permit scrutiny of those choices. Independent reproduction by an external group is still needed.

## Conclusions

Source-cohort discrimination did not establish stable performance across vascular endpoints and analysis pipelines. An isolated perfect ranking in eleven external samples coexisted with pipeline-dependent reversals, weak censored-event discrimination and unresolved observation dependence in a lesion-segment dataset. VascBridge-Instability provides public inputs, executable analyses and complete outputs for examining these limitations. It supports more careful development and independent testing of vascular research models, without claiming clinical risk prediction or treatment selection.

## Supporting information

Supplementary Data 1: analysis code, provenance and complete outputs

## Data availability

Public inputs are available in GEO under the seven accession records cited above. Supplementary Data 1 contains provenance and sample manifests, per-sample scores, all result tables and audit records. Public repository input files are retrieved from their source URLs rather than redistributed as new patient data. No access to private clinical records or proprietary datasets is required.

## Code availability

Supplementary Data 1 contains the executable analysis, public-input retrieval and figure code, environment lock, revision configuration and amendments. Its README documents the commands and limitations. The fixed seed and hashes identify the reported run. An independently maintained external replication has not yet been completed.

## Acknowledgments

We acknowledge the investigators and participants of the original public datasets and the GEO repository for making the source data available.

## Authors’ contributions

LY contributed to conceptualisation, methodology, formal analysis and writing of the original draft. YG contributed to methodology, validation and writing - review and editing. XZ, ZP and WW contributed to data curation, validation and writing – review and editing. LS contributed to supervision, methodology and writing - review and editing. JG contributed to conceptualisation, supervision, funding acquisition and writing - review and editing. The human authors take responsibility for the submitted work.

## Supplementary information

Supplementary Information contains Supplementary Table S1 and its explanatory note. Supplementary Data 1 contains the reproducibility materials: public-input retrieval, analysis and figure code; dependency lock; configuration and amendments; provenance and sample manifests; per-sample predictions and metric tables; and DOME-oriented reporting and internal consistency audits. The README documents the commands and limitations. Original processed matrices are retrieved from GEO.

