## Supplementary Information: Table S1 for "Cross study transfer of vascular transcriptomic classifiers in seven public cohorts"

**Supplementary Table S1. Source performance from one mean OOF score per sample**

| Model | AUROC | 95% interval | Average precision | Brier |
|---|---|---|---|---|
| L1 logistic | 0.947 | 0.869-0.996 | 0.915 | 0.092 |
| Elastic net | 0.949 | 0.873-0.998 | 0.897 | 0.130 |
| Linear SVM | 0.918 | 0.825-0.983 | 0.844 | Not applicable |
| Random forest | 0.911 | 0.812-0.981 | 0.773 | 0.131 |
| Prior only | 0.426 | 0.269-0.587 | 0.321 | 0.230 |

Intervals resample the 48 saved mean-OOF scores, conditional on this development exercise. They do not include training-set or hyperparameter-selection uncertainty.
