## Supplementary Data 1: analysis code, provenance and complete outputs for "Cross study transfer of vascular transcriptomic classifiers in seven public cohorts": audit_results.py

```
"""Read-only statistical checks plus explicitly labeled reporting corrections.

Run after benchmark_revision.py. Does not retrain or optimize any model.
The raw metrics are preserved. Manuscript-facing metrics suppress degenerate
bootstrap intervals, whose apparent precision is not sampling certainty.
"""
from pathlib import Path
import hashlib
import json
import sys
import importlib.metadata
import numpy as np
import pandas as pd
from sklearn.metrics import roc_auc_score
import benchmark_revision as b

HERE = Path(__file__).resolve().parent
OUT = HERE/'results'
metrics = pd.read_csv(OUT/'metrics.csv', float_precision='round_trip')
predictions = pd.read_csv(OUT/'predictions.csv', float_precision='round_trip')
source = pd.read_csv(OUT/'GSE98278_sample_manifest.csv')
raw = OUT/'metrics_before_reporting_audit.csv'
if not raw.exists():
    metrics.to_csv(raw, index=False)
else:
    metrics = pd.read_csv(raw, float_precision='round_trip')
metrics['bootstrap_degenerate'] = False
metrics['bootstrap_warning'] = ''
for i, row in metrics.iterrows():
    lo, hi = row.get('auroc_ci_low', np.nan), row.get('auroc_ci_high', np.nan)
    if np.isfinite(lo) and np.isfinite(hi) and lo == hi:
        metrics.loc[i, 'bootstrap_degenerate'] = True
        metrics.loc[i, 'bootstrap_warning'] = 'Nonparametric bootstrap cannot quantify uncertainty when every observed ordering is identical; interval suppressed; raw range retained in metrics_before_reporting_audit.csv.'
        metrics.loc[i, ['auroc_ci_low', 'auroc_ci_high']] = np.nan

# Distinguish restriction of full-source out-of-fold predictions from training
# separate, very small subset models. These are different estimands.
p = predictions[(predictions.variant == 'source_rank') & (predictions.model == 'L1_logistic') &
                (predictions.cohort == 'GSE98278') & (predictions.prediction_type == 'mean_repeated_nested_oof') &
                (predictions.subset == 'all')].merge(source[['sample_id', 'diameter_category']], on='sample_id', validate='one_to_one')
assert len(p) == 48
extra = []
for category in ['large', 'unknown']:
    q = p[p.diameter_category == category]
    row = b.metric_row('source_rank', 'L1_logistic', 'GSE98278', q.label.to_numpy(int), q.score.to_numpy(float), True,
                       'post-review descriptive within-category evaluation of full-source nested OOF scores; not subgroup-refitted model',
                       subset=f'full_source_oof_within_{category}')
    row['bootstrap_degenerate'] = False
    row['bootstrap_warning'] = 'Conditional-on-score sample interval; source donor independence unverified; subgroup models also depend on training size/mix.'
    extra.append(row)
metrics = pd.concat([metrics, pd.DataFrame(extra)], ignore_index=True)
metrics.to_csv(OUT/'metrics.csv', index=False)
pd.DataFrame(extra).to_csv(OUT/'full_source_oof_diameter_sensitivity.csv', index=False)

# Quantify active coefficients as well as the ANOVA-selected feature count.
coefs = pd.read_csv(OUT/'selected_features.csv')
missing = pd.read_csv(OUT/'missing_gene_audit.csv').fillna('')
auditrows = []
for _, row in missing.iterrows():
    if row.model == 'dummy_prior':
        continue
    c = coefs[(coefs.variant == row.variant) & (coefs.model == row.model)]
    active = c[np.abs(c.weight) > 1e-12]
    absent = set(str(row.missing_selected_genes).split('|')) - {''}
    active_missing = active[active.gene.isin(absent)]
    denom = active.weight.abs().sum()
    auditrows.append({'variant': row.variant, 'model': row.model, 'cohort': row.cohort,
                      'anova_selected_n': int(row.selected_n), 'nonzero_weight_n': len(active),
                      'nonzero_missing_n': len(active_missing),
                      'absolute_weight_missing_fraction': float(active_missing.weight.abs().sum()/denom) if denom else 0.0,
                      'note': 'RF uses nonzero feature importance; other models nonzero signed coefficient'})
pd.DataFrame(auditrows).to_csv(OUT/'active_feature_missingness.csv', index=False)

# Verify source OOF sample counts/disjointness and every reported metric against
# saved individual predictions; no outcome-driven model changes are allowed.
folds = pd.read_csv(OUT/'nested_folds.csv')
assert all(not (set(r.train_samples.split('|')) & set(r.test_samples.split('|'))) for _, r in folds.iterrows())
checks = []
for _, r in metrics.iterrows():
    if r.cohort != 'GSE98278' or r.subset != 'all' or r.variant == 'size_category_only':
        continue
    q = predictions[(predictions.variant == r.variant) & (predictions.model == r.model) &
                    (predictions.cohort == r.cohort) & (predictions.subset == 'all') &
                    (predictions.prediction_type == 'mean_repeated_nested_oof')]
    assert len(q) == 48 and q.sample_id.is_unique
    recomputed = roc_auc_score(q.label, q.score)
    assert abs(recomputed - r.auroc) < 1e-12
    f = predictions[(predictions.variant == r.variant) & (predictions.model == r.model) &
                    (predictions.cohort == r.cohort) & (predictions.subset == 'all') &
                    (predictions.prediction_type == 'nested_outer_fold')]
    assert set(f.groupby('sample_id').size()) == {3}
    checks.append({'variant': r.variant, 'model': r.model, 'auroc_recomputed': recomputed, 'unique_samples': 48,
                   'three_outer_predictions_per_sample': True, 'outer_fold_train_test_disjoint': True})

amendments = {
    'status': 'transparent post-review reporting and metadata audit, not preregistered',
    'sequence': [
        'Initial protocol and nested code were saved before revision model fits.',
        'Before new-cohort scores were inspected, public metadata audit replaced GSE41571 probe-median ratios with the official uncentered processed-signal supplement.',
        'Initial run completed main models and subgroup models but stopped at a keyword-only scikit-learn splitter call in size-only analysis.',
        'Splitter API corrected; sklearn 1.9 L1 ratio made explicit without changing effective penalty; PAD role and GSE41571 transformation prose corrected; entire analysis rerun successfully.',
        'After results were available, this reporting audit suppressed degenerate intervals, evaluated full-source OOF scores within diameter categories, and quantified active-weight missingness without fitting new models.'
    ],
    'dummy_oof_caution': 'Training-prevalence dummy has fold-specific constant scores; pooled mean-OOF AUROC may differ from 0.5 because fold prevalences vary. Every individual-fold dummy AUROC is 0.5; frozen external dummy AUROC is 0.5.',
    'source_log_caution': 'Cross-platform processed-signal scales are not biologically commensurate; signed-log sensitivity is a preprocessing stress diagnostic.',
    'PAD_caution': 'All deposited probe medians are zero across the PAD cohort; per-probe centering alters within-sample abundance ranks. This cohort cannot validate biological cross-bed transfer.',
    'diameter_caution': 'Within-stratum source-wide OOF and separately refit subgroup models answer different questions. Poor small-subset performance does not isolate causal diameter confounding.'
}
b.dump(HERE/'analysis_amendments.json', amendments)
b.dump(OUT/'result_audit.json', {'passed': True, 'source_checks': checks,
       'n_reported_rows': len(metrics), 'degenerate_auc_intervals_suppressed': int(metrics.bootstrap_degenerate.fillna(False).sum()),
       'full_source_oof_within_diameter': [{k: None if isinstance(v,float) and not np.isfinite(v) else v for k,v in r.items()} for r in extra]})
packages = ['numpy', 'pandas', 'scipy', 'scikit-learn', 'joblib', 'threadpoolctl']
(HERE/'requirements-lock.txt').write_text('\n'.join(f'{name}=={importlib.metadata.version(name)}' for name in packages)+'\n', encoding='utf-8')
b.dump(OUT/'reproducibility_manifest.json', {
    'python': sys.version, 'run_commands': ['python bmc_revision/benchmark_revision.py', 'python bmc_revision/audit_results.py'],
    'hashes': {str(p.relative_to(HERE)): b.sha(p) for p in [HERE/'benchmark_revision.py', HERE/'audit_results.py', HERE/'analysis_protocol.json', HERE/'analysis_amendments.json', HERE/'requirements-lock.txt'] + sorted(OUT.glob('*.csv')) + sorted(OUT.glob('*.json')) if p.name != 'reproducibility_manifest.json'}
})
print('Reporting audit complete. Full-source OOF by diameter:', [(x['subset'],x['auroc']) for x in extra])
```
