## Supplementary Data 1: analysis code, provenance and complete outputs for "Cross study transfer of vascular transcriptomic classifiers in seven public cohorts": benchmark_revision.py

```
"""Post-review public-data transfer benchmark. Run from any working directory.

No external outcome is used by model fitting, selection, scaling or tuning.
This revision is not a prospective or preregistered analysis: the first three
external cohorts were inspected in the preceding manuscript. All specified
models and sensitivities are retained irrespective of external discrimination.
"""
from __future__ import annotations

import argparse
import csv
import gzip
import hashlib
import io
import json
import os
import platform
import re
import sys
import time
import warnings
from pathlib import Path

for _v in ('OMP_NUM_THREADS', 'MKL_NUM_THREADS', 'OPENBLAS_NUM_THREADS'):
    os.environ.setdefault(_v, '1')
import joblib
import numpy as np
import pandas as pd
import scipy
from scipy.stats import rankdata
import sklearn
from sklearn.base import BaseEstimator, TransformerMixin
from sklearn.dummy import DummyClassifier
from sklearn.ensemble import RandomForestClassifier
from sklearn.exceptions import ConvergenceWarning
from sklearn.feature_selection import f_classif
from sklearn.linear_model import LogisticRegression
from sklearn.metrics import average_precision_score, brier_score_loss, roc_auc_score
from sklearn.model_selection import GridSearchCV, RepeatedStratifiedKFold, StratifiedKFold
from sklearn.pipeline import Pipeline
from sklearn.preprocessing import StandardScaler
from sklearn.svm import LinearSVC
from threadpoolctl import threadpool_limits

HERE = Path(__file__).resolve().parent
BASE = HERE.parent
DATA = HERE / 'data'
OUT = HERE / 'results'
SEED = 20261015
NBOOT = 2000
NPERM = 200
THREADS = 4
warnings.filterwarnings('ignore', category=FutureWarning)


def dump(path, obj):
    path.write_text(json.dumps(obj, indent=2, allow_nan=False, default=lambda o: o.item() if isinstance(o, np.generic) else str(o)), encoding='utf-8')


def sha(path):
    h = hashlib.sha256()
    with open(path, 'rb') as f:
        for b in iter(lambda: f.read(1024 * 1024), b''):
            h.update(b)
    return h.hexdigest()


def parse_series(path):
    lines = gzip.open(path, 'rt', errors='replace').read().splitlines()
    sm = {}
    for line in lines:
        if line.startswith('!Sample_'):
            row = next(csv.reader([line], delimiter='\t'))
            sm.setdefault(row[0][8:], []).append(row[1:])
    sm = {k: v if k == 'characteristics_ch1' else v[0] for k, v in sm.items()}
    begin = next(i for i, x in enumerate(lines) if x.startswith('!series_matrix_table_begin'))
    end = next(i for i in range(begin + 1, len(lines)) if lines[i].startswith('!series_matrix_table_end'))
    txt = '\n'.join(lines[begin + 1:end])
    expr = pd.read_csv(io.StringIO(txt), sep='\t', index_col=0) if txt.strip() else pd.DataFrame()
    return sm, expr.apply(pd.to_numeric, errors='coerce')


def strict_map(path):
    """Exclude ambiguous multi-symbol probes; do not silently select first gene."""
    result = {}
    with gzip.open(path, 'rt', errors='replace') as f:
        for line in f:
            if line.startswith('!platform_table_begin'):
                header = next(f).rstrip('\n').split('\t')
                symidx = header.index('Gene symbol')
                for line in f:
                    if line.startswith('!platform_table_end'):
                        break
                    vals = line.rstrip('\n').split('\t')
                    if len(vals) <= symidx:
                        continue
                    symbols = set(x.strip() for x in re.split(r'///| // ', vals[symidx]) if x.strip() not in ('', '---', 'NA'))
                    if len(symbols) == 1:
                        result[vals[0]] = symbols.pop()
                break
    return result


def map_genes(expr, mapping):
    valid = [x for x in expr.index if str(x) in mapping]
    mapped = expr.loc[valid].copy()
    mapped.index = [mapping[str(x)] for x in valid]
    return mapped.groupby(level=0, sort=True).median()


def load_cohorts():
    cohorts = {}
    platforms = {'GSE98278': 'GPL10558', 'GSE111782': 'GPL571', 'GSE27034': 'GPL570'}
    roles = {'GSE98278': 'source rupture versus elective stable AAA',
             'GSE111782': 'previously examined carotid symptomatic versus asymptomatic',
             'GSE198600': 'previously examined carotid recent symptomatic versus asymptomatic',
             'GSE27034': 'preprocessing-incompatible PAD stress control: deposited per-probe median-centered values do not preserve abundance ranks'}
    for accession, platform_id in platforms.items():
        sm, expr = parse_series(BASE / f'{accession}_series_matrix.txt.gz')
        expr = map_genes(expr, strict_map(BASE / f'{platform_id}.annot.gz'))
        ids = sm['geo_accession']
        if accession == 'GSE98278':
            groups = sm['characteristics_ch1'][1]
            y = np.array([int('ruptured' in s) for s in groups])
            assert all(('ruptured' in s) or ('elective stable' in s) for s in groups)
            size = ['intermediate' if 'intermediate' in s else 'large' if 'large' in s else 'unknown' for s in sm['characteristics_ch1'][2]]
            sample = pd.DataFrame({'sample_id': ids, 'label': y, 'diameter_category': size,
                                   'sample_title': sm['title'], 'donor_id': '',
                                   'donor_status': 'not deposited; sample independence cannot be verified'})
        elif accession == 'GSE111782':
            y = np.array([0 if 'asymptomatic' in s else 1 for s in sm['title']])
            assert all('symptomatic' in s for s in sm['title'])
            sample = pd.DataFrame({'sample_id': ids, 'label': y})
        else:
            y = np.array([int('PAD' in s) for s in sm['title']])
            assert all('PAD' in s or 'Control' in s for s in sm['title'])
            sample = pd.DataFrame({'sample_id': ids, 'label': y})
        cohorts[accession] = {'expression': expr.loc[:, ids], 'samples': sample, 'role': roles[accession]}
    sm, _ = parse_series(BASE / 'GSE198600_series_matrix.txt.gz')
    ids = sm['geo_accession']
    y = np.array([0 if 'Asymp' in s else 1 for s in sm['title']])
    assert all('Asymp' in s or 'Symp' in s for s in sm['title'])
    expr = pd.read_csv(BASE / 'GSE198600_fpkm.tsv', sep='\t', index_col=0)
    assert set(ids) == set(expr.columns)
    cohorts['GSE198600'] = {'expression': expr.loc[:, ids], 'samples': pd.DataFrame({'sample_id': ids, 'label': y}), 'role': roles['GSE198600']}
    for accession in ('GSE41571', 'GSE163154', 'GSE21545'):
        expr = pd.read_csv(DATA / f'{accession}_expression.tsv.gz', sep='\t', index_col=0)
        sample = pd.read_csv(DATA / f'{accession}_samples.csv').fillna('')
        assert expr.index.is_unique and sample.sample_id.is_unique
        assert set(expr.columns) == set(sample.sample_id)
        cohorts[accession] = {'expression': expr.loc[:, sample.sample_id], 'samples': sample,
                             'role': str(sample.endpoint_role.iloc[0])}
    for accession, d in cohorts.items():
        assert d['expression'].index.is_unique
        assert (d['expression'].columns.to_numpy() == d['samples'].sample_id.to_numpy()).all()
        d['samples'].to_csv(OUT / f'{accession}_sample_manifest.csv', index=False)
    return cohorts


def transform_cohort(expr, genes, mode, accession):
    a = expr.reindex(genes).T.to_numpy(dtype=float)
    if mode.endswith('rank'):
        finite = np.isfinite(a)
        ranks = rankdata(a, axis=1, method='average', nan_policy='omit')
        n = finite.sum(axis=1)
        a = (ranks - 1) / np.maximum(n[:, None] - 1, 1)
    elif mode == 'source_log':
        if accession == 'GSE98278':
            a = np.sign(a) * np.log2(1 + np.abs(a))
        elif accession in ('GSE198600', 'GSE41571'):
            a = np.log2(1 + np.maximum(a, 0))
        # Other cohorts are already deposited on log/ratio scales.  They are
        # intentionally not re-centered using external cohort averages.
    else:
        raise ValueError(mode)
    return a


class TrainOnlySelect(BaseEstimator, TransformerMixin):
    """Impute, variance-filter, and select supervised features using fit rows only."""
    def __init__(self, k=100, variance_limit=5000):
        self.k = k
        self.variance_limit = variance_limit

    def fit(self, X, y):
        X = np.asarray(X, dtype=float)
        with warnings.catch_warnings():
            warnings.simplefilter('ignore', RuntimeWarning)
            self.median_ = np.nanmedian(X, axis=0)
        # Source genes missing in all training samples cannot be selected.
        valid = np.isfinite(self.median_)
        self.median_[~valid] = 0.0
        z = np.where(np.isfinite(X), X, self.median_)
        var = np.var(z, axis=0)
        valid &= var > 1e-14
        idx = np.flatnonzero(valid)
        idx = idx[np.argsort(-var[idx], kind='stable')[:self.variance_limit]]
        if len(idx) == 0:
            raise ValueError('No nonconstant training features')
        with warnings.catch_warnings():
            warnings.simplefilter('ignore', RuntimeWarning)
            f, _ = f_classif(z[:, idx], y)
        f = np.nan_to_num(f, nan=-np.inf)
        self.indices_ = idx[np.argsort(-f, kind='stable')[:min(self.k, len(idx))]]
        return self

    def transform(self, X):
        z = np.asarray(X, dtype=float)[:, self.indices_]
        return np.where(np.isfinite(z), z, self.median_[self.indices_])


def make_pipeline(name):
    if name == 'L1_logistic':
        clf = LogisticRegression(penalty='l1', l1_ratio=1.0, solver='liblinear', C=0.1, max_iter=10000, random_state=SEED)
        grid = {'select__k': [25, 100], 'model__C': [0.1, 1.0]}
    elif name == 'elastic_net':
        clf = LogisticRegression(penalty='elasticnet', solver='saga', l1_ratio=0.5, C=0.1, max_iter=10000, tol=1e-4, random_state=SEED)
        grid = {'select__k': [25, 100], 'model__C': [0.1, 1.0]}
    elif name == 'linear_SVM':
        clf = LinearSVC(C=0.1, max_iter=20000, random_state=SEED, dual='auto')
        grid = {'select__k': [25, 100], 'model__C': [0.1, 1.0]}
    elif name == 'random_forest':
        clf = RandomForestClassifier(n_estimators=200, max_features='sqrt', min_samples_leaf=2, random_state=SEED, n_jobs=1)
        grid = {'select__k': [25, 100], 'model__max_depth': [3, None]}
    elif name == 'dummy_prior':
        return Pipeline([('model', DummyClassifier(strategy='prior'))]), {}
    else:
        raise ValueError(name)
    return Pipeline([('select', TrainOnlySelect()), ('scale', StandardScaler()), ('model', clf)]), grid


def tune(name, X, y, seed=SEED):
    pipe, grid = make_pipeline(name)
    if name == 'dummy_prior':
        pipe.fit(np.nan_to_num(X), y)
        return pipe, {}, None
    inner = StratifiedKFold(3, shuffle=True, random_state=seed)
    search = GridSearchCV(pipe, grid, scoring='roc_auc', cv=inner, refit=True, n_jobs=1, error_score='raise', return_train_score=False)
    search.fit(X, y)
    return search.best_estimator_, search.best_params_, float(search.best_score_)


def predict(model, X):
    if hasattr(model, 'predict_proba'):
        return model.predict_proba(X)[:, 1], True
    return model.decision_function(X), False


def bootstrap_auc(y, score, seed):
    """Stratified resampling of source samples; conditional on fitted scores."""
    rng = np.random.default_rng(seed)
    neg, pos = score[y == 0], score[y == 1]
    n, p = len(neg), len(pos)
    a = pos[rng.integers(0, p, (NBOOT, p))]
    b = neg[rng.integers(0, n, (NBOOT, n))]
    auc = ((a[:, :, None] > b[:, None, :]).sum((1, 2)) + 0.5 * (a[:, :, None] == b[:, None, :]).sum((1, 2))) / (n * p)
    return [float(np.quantile(auc, 0.025)), float(np.quantile(auc, 0.975))]


def harrell_c(event, times, risk):
    comparable = (times[:, None] < times[None, :]) & event.astype(bool)[:, None]
    num = comparable.sum()
    if not num:
        return np.nan
    correct = (risk[:, None] > risk[None, :]) + 0.5 * (risk[:, None] == risk[None, :])
    return float(np.sum(correct * comparable) / num)


def bootstrap_c(event, times, risk, seed):
    rng = np.random.default_rng(seed)
    vals = []
    for _ in range(NBOOT):
        idx = rng.integers(0, len(event), len(event))
        c = harrell_c(event[idx], times[idx], risk[idx])
        if np.isfinite(c):
            vals.append(c)
    return [float(np.quantile(vals, 0.025)), float(np.quantile(vals, 0.975))]


def metric_row(variant, model, cohort, y, p, probability, role, ci=True, subset='all', seed=SEED):
    row = {'variant': variant, 'model': model, 'cohort': cohort, 'subset': subset,
           'role': role, 'n': len(y), 'positive': int(sum(y)), 'negative': int(sum(y == 0)),
           'auroc': float(roc_auc_score(y, p)), 'auprc': float(average_precision_score(y, p)),
           'prevalence': float(np.mean(y)), 'brier': float(brier_score_loss(y, p)) if probability else np.nan,
           'auroc_ci_low': np.nan, 'auroc_ci_high': np.nan, 'harrell_c': np.nan,
           'c_ci_low': np.nan, 'c_ci_high': np.nan,
           'ci_scope': 'stratified sample bootstrap conditional on scores; excludes model-selection uncertainty' if ci else 'not estimated: unresolved repeated-donor dependence'}
    if ci:
        row['auroc_ci_low'], row['auroc_ci_high'] = bootstrap_auc(y, p, seed)
    return row


def prediction_rows(variant, model, cohort, sample, scores, kind, **extra):
    rows = []
    for (_, s), p in zip(sample.iterrows(), scores):
        rows.append({'variant': variant, 'model': model, 'cohort': cohort,
                     'sample_id': s.sample_id, 'label': int(s.label), 'score': float(p),
                     'prediction_type': kind, 'donor_id': s.get('donor_id', ''),
                     'followup_days': s.get('followup_days', ''), **extra})
    return rows


def nested_cv(variant, name, X, y, sample, subset='all'):
    cv = RepeatedStratifiedKFold(n_splits=4, n_repeats=3, random_state=SEED)
    sums, counts = np.zeros(len(y)), np.zeros(len(y), dtype=int)
    predictions, folds = [], []
    for fi, (train, test) in enumerate(cv.split(X, y)):
        model, params, inner_auc = tune(name, X[train], y[train], SEED + fi)
        p, probability = predict(model, X[test])
        sums[test] += p
        counts[test] += 1
        folds.append({'variant': variant, 'model': name, 'subset': subset, 'fold': fi,
                      'repeat': fi // 4, 'outer_auroc': roc_auc_score(y[test], p),
                      'n_train': len(train), 'n_test': len(test), 'inner_auroc': inner_auc,
                      'selected_params': json.dumps(params, sort_keys=True),
                      'train_samples': '|'.join(sample.iloc[train].sample_id),
                      'test_samples': '|'.join(sample.iloc[test].sample_id)})
        predictions += prediction_rows(variant, name, 'GSE98278', sample.iloc[test], p, 'nested_outer_fold', repeat=fi // 4, fold=fi, subset=subset)
    assert np.all(counts == 3)
    mean_p = sums / counts
    predictions += prediction_rows(variant, name, 'GSE98278', sample, mean_p, 'mean_repeated_nested_oof', subset=subset)
    metric = metric_row(variant, name, 'GSE98278', y, mean_p, probability,
                        'nested 4-fold CV repeated 3 times; unique-sample mean OOF', subset=subset)
    metric['fold_auroc_mean'] = float(np.mean([z['outer_auroc'] for z in folds]))
    metric['fold_auroc_sd'] = float(np.std([z['outer_auroc'] for z in folds], ddof=1))
    return metric, predictions, folds


def fixed_oof(X, y, sample, keep_predictions=False):
    sums = np.zeros(len(y))
    counts = np.zeros(len(y), dtype=int)
    rows = []
    cv = RepeatedStratifiedKFold(n_splits=4, n_repeats=3, random_state=SEED)
    for fi, (train, test) in enumerate(cv.split(X, y)):
        pipe, _ = make_pipeline('L1_logistic')
        pipe.set_params(select__k=100, model__C=0.25)
        pipe.fit(X[train], y[train])
        p, _ = predict(pipe, X[test])
        sums[test] += p
        counts[test] += 1
        if keep_predictions:
            s = sample.iloc[test].copy()
            s['label'] = y[test]
            rows += prediction_rows('source_rank_fixed_permutation', 'L1_logistic', 'GSE98278', s, p, 'fixed_baseline_oof', repeat=fi // 4, fold=fi)
    mean_p = sums / counts
    return float(roc_auc_score(y, mean_p)), mean_p, rows


def run_permutation(X, y, sample):
    print('Starting 200 whole-pipeline fixed-baseline permutations', flush=True)
    observed, p, rows = fixed_oof(X, y, sample, True)
    rng = np.random.default_rng(SEED + 10000)
    labels = [rng.permutation(y) for _ in range(NPERM)]
    def one(i):
        auc, predictions, _ = fixed_oof(X, labels[i], sample)
        return i, auc, predictions
    # Threads share the immutable source matrix; each fit owns its own pipeline.
    result = joblib.Parallel(n_jobs=THREADS, prefer='threads', verbose=5)(joblib.delayed(one)(i) for i in range(NPERM))
    null = np.array([r[1] for r in result])
    out = []
    for i, auc, predictions in result:
        for sid, label, val in zip(sample.sample_id, labels[i], predictions):
            out.append({'permutation': i, 'sample_id': sid, 'permuted_label': int(label), 'mean_repeated_oof_score': float(val), 'permutation_auroc': auc})
    pd.DataFrame(out).to_csv(OUT / 'permutation_sample_predictions.csv.gz', index=False)
    pd.DataFrame({'permutation': np.arange(NPERM), 'auroc': null}).to_csv(OUT / 'permutation_null.csv', index=False)
    pd.DataFrame(rows).to_csv(OUT / 'permutation_observed_fold_predictions.csv', index=False)
    summary = {'n_permutations': NPERM, 'observed_auroc': observed, 'null_mean': float(null.mean()),
               'null_percentiles_2_5_97_5': [float(x) for x in np.quantile(null, [0.025, 0.975])],
               'one_sided_empirical_p': float((1 + np.sum(null >= observed)) / (1 + NPERM)),
               'fixed_hyperparameters': {'k': 100, 'C': 0.25, 'variance_limit': 5000},
               'statistic': 'unique-sample AUROC of mean predictions from 4-fold CV repeated 3 times',
               'splits': 'stratified splits regenerated under each permuted label vector using the same fixed splitter seed and 3 repeats as the observed analysis',
               'scope': 'fixed baseline only, not a significance test of the separately hyperparameter-tuned models',
               'label_exchangeability_limitation': 'unrestricted sample permutations; donor IDs not deposited; does not address diameter confounding or post-review selection'}
    dump(OUT / 'permutation_summary.json', summary)
    return summary


def build_protocol():
    return {'date': '2026-10-15', 'analysis_status': 'post-review revision; not a preregistration',
            'prior_external_inspection': ['GSE111782', 'GSE198600', 'GSE27034'],
            'new_outcome_definitions_fixed_before_prediction': ['GSE41571 rupture-associated macrophage plaques', 'GSE163154 paired plaque IPH, segment-descriptive only', 'GSE21545 future ischemic event with censoring, full and 30-day landmark'],
            'primary': {'source': 'GSE98278', 'feature_vocabulary': 'all unambiguous annotated genes present in source alone',
                        'normalization': 'within-sample ranks of observed source-vocabulary genes; missing entries remain missing',
                        'missing_gene_policy': 'impute training median, learned inside each training fold; never replace absent measured genes by zero expression',
                        'variance_limit': 5000, 'supervised_k_grid': [25, 100],
                        'models': ['L1_logistic', 'elastic_net', 'linear_SVM', 'random_forest', 'dummy_prior'],
                        'C_grid': [0.1, 1.0], 'elastic_net_l1_ratio': 0.5,
                        'forest': {'trees': 200, 'min_samples_leaf': 2, 'max_depth_grid': [3, None], 'max_features': 'sqrt'},
                        'outer_cv': '4 folds x 3 repeats, sample stratified', 'inner_cv': '3 folds, source-only AUROC selection',
                        'primary_internal_unit': 'one mean OOF score per source sample',
                        'external_models': 'refit each source-tuned model to all source samples; no external calibration, feature/outcome selection, or threshold search'},
            'sensitivities': {'common_rank': 'unambiguous gene intersection of source plus all three previously examined external cohorts; explicitly transductive label-blind availability; L1 only',
                              'source_log': 'source-vocabulary signed log2(1+abs intensity) in background-corrected AAA; log2(1+FPKM) for RNA-seq and log2(1+uncentered processed intensity) for GSE41571 official supplementary raw columns; other processed log matrices retained; source-fitted scaling; L1 only; cross-platform scale-mismatch diagnostic',
                              'size': 'nested source L1 within large (16) and unknown (17) size strata plus category-only 3-repeat OOF baseline; intermediate stratum has no ruptures'},
            'uncertainty': {'bootstrap': '2000 stratified sample resamples for AUROC, conditional on scores; excludes training and model-selection uncertainty',
                            'GSE163154': 'no patient CI because 43 paired/clustered segments from 24 original participants lack donor mapping',
                            'GSE21545': 'Harrell C-index and 2000 unique-donor bootstrap resamples; no binary classification of censored subjects as event-free; 30-day landmark pre-specified'},
            'permutation': {'n': NPERM, 'fixed_baseline': 'source-rank L1, k=100, C=0.25; complete train-only selection refit each fold',
                            'splits': 'same 4-fold x 3-repeat label-stratified splitter rule for observed/null',
                            'p': '(1 + null AUROC >= observed)/(1 + 200)', 'tuned_model_test': False},
            'mapping': 'public GEO annotations; unambiguous gene symbols only, within-gene probe median; no first-symbol choice for multi-gene probes',
            'no_optimization_on_external': True, 'seed': SEED}


def run():
    OUT.mkdir(exist_ok=True, parents=True)
    # Protocol is materialized before this revision computes any outcomes.
    protocol_path = HERE / 'analysis_protocol.json'
    dump(protocol_path, build_protocol())
    dump(OUT / 'environment.json', {'python': sys.version, 'platform': platform.platform(), 'numpy': np.__version__,
         'pandas': pd.__version__, 'scipy': scipy.__version__, 'scikit_learn': sklearn.__version__, 'joblib': joblib.__version__,
         'command': 'python bmc_revision/benchmark_revision.py', 'threads': THREADS})
    cohorts = load_cohorts()
    source = cohorts['GSE98278']
    y = source['samples'].label.to_numpy(int)
    source_genes = sorted(source['expression'].index)
    common_genes = sorted(set(source_genes).intersection(*(set(cohorts[g]['expression'].index) for g in ['GSE111782', 'GSE198600', 'GSE27034'])))
    genesets = {'source_rank': source_genes, 'common_rank': common_genes, 'source_log': source_genes}
    dump(OUT / 'gene_vocabulary.json', genesets)
    print(f'Loaded source n={len(y)}, source genes={len(source_genes)}, common genes={len(common_genes)}', flush=True)
    metrics, all_predictions, foldrows, featurerows, missingrows = [], [], [], [], []
    variants = [('source_rank', ['L1_logistic', 'elastic_net', 'linear_SVM', 'random_forest', 'dummy_prior']),
                ('common_rank', ['L1_logistic']), ('source_log', ['L1_logistic'])]
    final_params = {}
    def save_tables():
        pd.DataFrame(metrics).to_csv(OUT / 'metrics.csv', index=False)
        pd.DataFrame(all_predictions).to_csv(OUT / 'predictions.csv', index=False)
        pd.DataFrame(foldrows).to_csv(OUT / 'nested_folds.csv', index=False)
        pd.DataFrame(featurerows).to_csv(OUT / 'selected_features.csv', index=False)
        pd.DataFrame(missingrows).to_csv(OUT / 'missing_gene_audit.csv', index=False)
        dump(OUT / 'final_source_tuning.json', final_params)
    for variant, names in variants:
        genes = genesets[variant]
        matrices = {g: transform_cohort(d['expression'], genes, variant, g) for g, d in cohorts.items()}
        X = matrices['GSE98278']
        for name in names:
            start = time.monotonic()
            print(f'Nested CV {variant}/{name}', flush=True)
            row, preds, folds = nested_cv(variant, name, X, y, source['samples'])
            metrics.append(row); all_predictions += preds; foldrows += folds
            model, params, inner_auc = tune(name, X, y)
            final_params[f'{variant}/{name}'] = {'parameters': params, 'source_inner_auroc': inner_auc}
            joblib.dump(model, OUT / f'frozen_{variant}_{name}.joblib')
            selected = model.named_steps['select'].indices_ if name != 'dummy_prior' else np.array([], dtype=int)
            classifier = model.named_steps['model']
            weights = classifier.coef_[0] if hasattr(classifier, 'coef_') else classifier.feature_importances_ if hasattr(classifier, 'feature_importances_') else np.zeros(len(selected))
            for idx, weight in zip(selected, weights):
                featurerows.append({'variant': variant, 'model': name, 'gene': genes[idx], 'weight': float(weight)})
            for accession, d in cohorts.items():
                if accession == 'GSE98278':
                    continue
                mat = matrices[accession]
                p, probability = predict(model, mat)
                sy = d['samples'].label.to_numpy(int)
                all_predictions += prediction_rows(variant, name, accession, d['samples'], p, 'frozen_external')
                missing = [genes[i] for i in selected if genes[i] not in d['expression'].index]
                missingrows.append({'variant': variant, 'model': name, 'cohort': accession, 'vocabulary_n': len(genes),
                                    'available_vocabulary_n': len(set(genes) & set(d['expression'].index)),
                                    'selected_n': len(selected), 'selected_missing_n': len(missing),
                                    'selected_missing_fraction': len(missing)/len(selected) if len(selected) else 0,
                                    'missing_selected_genes': '|'.join(missing),
                                    'finite_selected_values_fraction': float(np.isfinite(mat[:, selected]).mean()) if len(selected) else 1})
                if accession == 'GSE21545':
                    times = d['samples'].followup_days.to_numpy(float)
                    for subset, mask in [('all', np.ones(len(times), dtype=bool)), ('30_day_landmark', times > 30)]:
                        t, e, score = times[mask], sy[mask], p[mask]
                        if subset != 'all': t = t - 30
                        c = harrell_c(e, t, score)
                        low, high = bootstrap_c(e, t, score, SEED)
                        metrics.append({'variant': variant, 'model': name, 'cohort': accession, 'subset': subset,
                                        'role': d['role'], 'n': len(t), 'positive': int(e.sum()), 'negative': int(sum(e == 0)),
                                        'auroc': np.nan, 'auprc': np.nan, 'prevalence': np.nan, 'brier': np.nan,
                                        'harrell_c': c, 'c_ci_low': low, 'c_ci_high': high,
                                        'ci_scope': '2000 individual bootstrap samples; conditional on scores; Harrell C affected by censoring distribution'})
                else:
                    metrics.append(metric_row(variant, name, accession, sy, p, probability, d['role'], ci=accession != 'GSE163154'))
            save_tables()
            print(f'Complete {variant}/{name}: nested AUROC={row["auroc"]:.3f}, elapsed={time.monotonic()-start:.1f}s', flush=True)
    X = transform_cohort(source['expression'], source_genes, 'source_rank', 'GSE98278')
    for size in ('large', 'unknown'):
        mask = source['samples'].diameter_category.to_numpy() == size
        row, preds, folds = nested_cv('source_rank', 'L1_logistic', X[mask], y[mask], source['samples'].loc[mask].reset_index(drop=True), subset=f'diameter_{size}')
        metrics.append(row); all_predictions += preds; foldrows += folds
        save_tables()
        print(f'Within-size nested analysis {size}: n={sum(mask)}, AUROC={row["auroc"]:.3f}', flush=True)
    # Negative control for publicly observed size categories (unknown is a
    # category, not a fabricated numeric diameter or imputed measurement).
    sizeX = pd.get_dummies(source['samples'].diameter_category).to_numpy(float)
    cv = RepeatedStratifiedKFold(n_splits=4, n_repeats=3, random_state=SEED)
    sums = np.zeros(len(y)); counts = np.zeros(len(y))
    for fi, (train, test) in enumerate(cv.split(sizeX, y)):
        q = LogisticRegression(C=1.0, solver='liblinear', random_state=SEED).fit(sizeX[train], y[train])
        p = q.predict_proba(sizeX[test])[:, 1]
        sums[test] += p; counts[test] += 1
        all_predictions += prediction_rows('size_category_only', 'L2_logistic_fixed_C1', 'GSE98278', source['samples'].iloc[test], p, 'fixed_covariate_oof', repeat=fi//4, fold=fi)
    avg = sums/counts
    metrics.append(metric_row('size_category_only', 'L2_logistic_fixed_C1', 'GSE98278', y, avg, True, 'public diameter category only; unknown retained as category'))
    all_predictions += prediction_rows('size_category_only', 'L2_logistic_fixed_C1', 'GSE98278', source['samples'], avg, 'mean_repeated_oof')
    pd.crosstab(source['samples'].diameter_category, source['samples'].label).to_csv(OUT / 'diameter_label_counts.csv')
    save_tables()
    permutation = run_permutation(X, y, source['samples'])
    audit = {'completed': True, 'protocol_sha256': sha(protocol_path), 'script_sha256': sha(Path(__file__)),
             'source_n': len(y), 'source_genes': len(source_genes), 'common_genes': len(common_genes),
             'primary_models': variants[0][1], 'n_metrics': len(metrics), 'n_prediction_rows': len(all_predictions),
             'source_metadata_limitation': 'no donor identifiers; cross-validation and source CIs are sample-level, not verified patient-independent',
             'external_reuse': 'first three external cohorts are previously examined, not untouched confirmatory tests',
             'permutation': permutation,
             'input_sha256': {str(p.relative_to(BASE)): sha(p) for p in list(BASE.glob('*series_matrix.txt.gz')) + list(BASE.glob('*.annot.gz')) + [BASE/'GSE198600_fpkm.tsv'] + list(DATA.glob('*_expression.tsv.gz')) + list(DATA.glob('*_samples.csv'))}}
    dump(OUT / 'run_summary.json', audit)
    print('COMPLETE', flush=True)


if __name__ == '__main__':
    with threadpool_limits(limits=1):
        run()
```
