## Supplementary Data 1: analysis code, provenance and complete outputs for "Cross study transfer of vascular transcriptomic classifiers in seven public cohorts": build_reproducibility_package.py

```
"""Build and verify the small, portable Additional file 1 code/results archive.

Only explicit public-science files are packaged. Large expression sources and
private manuscript administrative material are excluded. Science scripts and
results are byte-identical; packaging-only audit edits are fully documented.
"""
from pathlib import Path
import csv
import hashlib
import io
import json
import re
import zipfile

HERE = Path(__file__).resolve().parent
SUBMISSION = HERE / 'submission'
OUTPUT = SUBMISSION / 'Additional_file_1_reproducibility.zip'
FILES = [
    'download_public_inputs.py', 'public_input_snapshot.json', 'cohort_extension.py',
    'benchmark_revision.py', 'audit_results.py', 'analysis_protocol.json',
    'analysis_amendments.json', 'requirements-lock.txt', 'requirements-reporting.txt',
    'make_revision_figures.py', 'README_REPRODUCE.txt', 'build_reproducibility_package.py',
    'cohort_audit.json', 'cohort_audit.csv', 'cohort_audit.txt',
    'independent_data_review.json', 'independent_data_review.txt',
    'editorial_result_audit.json', 'editorial_result_review.py', 'dome_reporting_audit.csv',
    'public_input_verification.json', 'acquisition_reconstruction_test.json'
]
DATA_FILES = ['extension_samples_all.csv'] + [
    f'{accession}_{suffix}' for accession in ['GSE41571', 'GSE21545', 'GSE163154']
    for suffix in ['samples.csv', 'series_metadata.json']
]


def digest(data):
    return hashlib.sha256(data).hexdigest()


def json_bytes(data):
    return (json.dumps(data, indent=2, ensure_ascii=False)+'\n').encode('utf-8')


def portable_paths(value):
    if isinstance(value, dict):
        return {k: portable_paths(v) for k, v in value.items()}
    if isinstance(value, list):
        return [portable_paths(v) for v in value]
    if isinstance(value, str):
        if re.match(r'^[A-Za-z]:[\\/]', value):
            path = value.replace('\\', '/')
            if '/bmc_revision/' in path:
                return path.split('/bmc_revision/', 1)[1]
            return '../' + path.rsplit('/', 1)[-1]
        if value.startswith('data\\'):
            return value.replace('\\', '/')
    return value


def package_copy(name, original):
    """Clean distribution copies only; never change original audit/results."""
    edits = []
    output = original
    if name == 'cohort_audit.json':
        data = portable_paths(json.loads(original))
        data['mapping_difference_from_original'] = (
            'The original exploratory run chose the first symbol for multi-symbol probes. '
            'The revision source and extension pipelines both exclude ambiguous multi-symbol probes. '
            'GPL570 retains 42,904/54,675 probe rows mapping to 20,848 genes; GPL6104 retains '
            '20,502/22,185 probe rows mapping to 17,184 genes. The common mapping rule was '
            'independently checked against the revision engine.')
        output = json_bytes(data)
        edits += ['Replace local absolute annotation paths by relative paths.',
                  'Replace superseded mapping-change preparation instruction with the verified implemented rule.']
    elif name == 'independent_data_review.json':
        data = json.loads(original)
        data['review_scope'] = ('Initial cohort metadata were audited before inspecting extension-model outcomes; '
                               'the completed review independently recomputed the saved final metrics, folds and predictions.')
        data.pop('critical_findings', None)
        data['code_review']['nonblocking_notes'] = [x for x in data['code_review']['nonblocking_notes'] if 'penalty=l1/l1_ratio0' not in x]
        data['final_scientific_limitations'] = [x.replace('; no supported70percent journal acceptance probability.', '.') for x in data['final_scientific_limitations']]
        output = json_bytes(data)
        edits += ['Retain final numerical review; remove superseded intermediate correction requests and resolved sklearn warning.',
                  'Clarify initial metadata audit versus completed results audit; omit journal-acceptance discussion.']
    elif name == 'independent_data_review.txt':
        output = original.decode('utf-8').replace('; no supported70percent journal acceptance probability.', '.').encode('utf-8')
        edits += ['Remove journal-acceptance discussion from scientific supplementary audit.']
    elif name == 'dome_reporting_audit.csv':
        data = original.decode('utf-8')
        data = data.replace('Public metadata and biological relevance described in narrative_sections.json',
                            'Public metadata and biological relevance in cohort_audit.json and analysis_protocol.json')
        data = data.replace('Local evidence ready,Public permanent archive still requires actual deposition',
                            'Supplied in Additional file 1,No permanent repository identifier is assigned by this ZIP archive')
        output = data.encode('utf-8')
        edits += ['Point reporting evidence to included files; replace obsolete local-preparation status with supplementary-archive status.']
    if output == original:
        edits = []
    return output, edits


def build():
    SUBMISSION.mkdir(exist_ok=True)
    payloads, edits = {}, []
    for name in FILES:
        source = HERE/name
        original = source.read_bytes()
        content, changes = package_copy(name, original)
        payloads['bmc_revision/'+name] = content
        if changes:
            edits.append({'file': name, 'source_sha256': digest(original), 'distributed_sha256': digest(content), 'changes': changes})
    for name in DATA_FILES:
        payloads['bmc_revision/data/'+name] = (HERE/'data'/name).read_bytes()
    result_files = sorted((HERE/'results').iterdir())
    for path in result_files:
        if path.is_file():
            if path.stat().st_size > 5*1024*1024:
                raise ValueError(f'Unexpectedly large result file: {path.name}')
            if path.suffix not in {'.csv', '.json', '.gz', '.joblib'}:
                raise ValueError(f'Unreviewed result type: {path.name}')
            payloads['bmc_revision/results/'+path.name] = path.read_bytes()
    notes = {
        'title': 'Additional file 1: reproducible public-data benchmark code, numerical results and audits',
        'archive_layout': 'Keep the bmc_revision/ folder; run commands from its parent.',
        'scientific_scripts_and_results': 'Included without changing their bytes or numerical values.',
        'public_inputs': 'Large GEO expression/annotation archives excluded; download_public_inputs.py acquires official sources using public_input_snapshot.json.',
        'packaging_only_changes': edits,
        'historical_manifests': 'Historical run manifests record original-run hashes; MANIFEST_SHA256.json describes the distributed package. Platform line endings and excluded unused local public files are explained in README_REPRODUCE.txt.',
        'internal_review': 'Computational/internal audit, not external peer review.',
        'license_note': 'This archive assigns no new license to third-party public source data; use the official source terms.'
    }
    payloads['bmc_revision/PACKAGE_NOTES.json'] = json_bytes(notes)
    manifest = {'algorithm': 'SHA-256', 'scope': 'Every archive member except this manifest itself',
                'files': {name: {'bytes':len(data), 'sha256':digest(data)} for name,data in sorted(payloads.items())}}
    payloads['bmc_revision/MANIFEST_SHA256.json'] = json_bytes(manifest)
    for name, data in payloads.items():
        if name.endswith(('.py','.json','.txt','.csv')):
            text = data.decode('utf-8')
            if re.search(r'(?i)(?:\b[a-z]:[\\/]|\.codex[\\/])', text):
                raise ValueError(f'Local/private path marker in {name}')
    with zipfile.ZipFile(OUTPUT, 'w', compression=zipfile.ZIP_DEFLATED, compresslevel=9) as archive:
        for name, data in sorted(payloads.items()):
            entry = zipfile.ZipInfo(name, date_time=(2026,10,15,0,0,0))
            entry.compress_type = zipfile.ZIP_DEFLATED
            entry.external_attr = 0o644 << 16
            archive.writestr(entry, data, compress_type=zipfile.ZIP_DEFLATED, compresslevel=9)
    if OUTPUT.stat().st_size >= 20*1024*1024:
        raise ValueError('Additional file exceeds 20 MiB')
    with zipfile.ZipFile(OUTPUT) as archive:
        if archive.testzip() is not None:
            raise ValueError('ZIP CRC check failed')
        for name in archive.namelist():
            if not name.startswith('bmc_revision/') or '..' in Path(name).parts or name.startswith('/'):
                raise ValueError(f'Unsafe/nonportable archive path: {name}')
        packaged_manifest = json.loads(archive.read('bmc_revision/MANIFEST_SHA256.json'))
        for name, item in packaged_manifest['files'].items():
            data = archive.read(name)
            assert len(data) == item['bytes'] and digest(data) == item['sha256'], name
        for path in result_files:
            if path.is_file():
                assert archive.read('bmc_revision/results/'+path.name) == path.read_bytes(), path.name
        for name in ['benchmark_revision.py','audit_results.py','cohort_extension.py','download_public_inputs.py','requirements-lock.txt','make_revision_figures.py']:
            assert archive.read('bmc_revision/'+name) == (HERE/name).read_bytes(), name
        metrics = list(csv.DictReader(io.StringIO(archive.read('bmc_revision/results/metrics.csv').decode())))
        assert len(metrics) == 61
    report = {'archive': OUTPUT.name, 'bytes': OUTPUT.stat().st_size, 'mib': OUTPUT.stat().st_size/1024**2,
              'sha256': digest(OUTPUT.read_bytes()), 'file_count': len(payloads), 'crc_valid': True,
              'manifest_hashes_valid': True, 'portable_bmc_revision_layout': True,
              'science_results_byte_identical': True, 'local_private_path_scan_passed': True,
              'large_expression_inputs_excluded': True, 'metric_rows': len(metrics),
              'packaging_audit_copies_changed': [x['file'] for x in edits]}
    (SUBMISSION/'Additional_file_1_validation.json').write_bytes(json_bytes(report))
    print(json.dumps(report,indent=2))


if __name__ == '__main__':
    build()
```
