## Supplementary Data 1: analysis code, provenance and complete outputs for "Cross study transfer of vascular transcriptomic classifiers in seven public cohorts": cohort_extension.py

```
"""Acquire and audit public GEO extension cohorts without inspecting model scores.

This script writes gene-by-GSM processed expression matrices and explicit metadata.
No endpoint is silently treated as equivalent to source AAA rupture. In particular,
BiKE follow-up event status is not symptomatic status and IPH segments are not
assumed to represent independent donors.
"""
from pathlib import Path
import csv
import gzip
import hashlib
import io
import json
import re
import urllib.request
from datetime import datetime, timezone

import numpy as np
import pandas as pd

BASE = Path(__file__).resolve().parent
DATA = BASE / 'data'
SOURCE = BASE.parent
DATA.mkdir(exist_ok=True)


def sha256(path):
    h = hashlib.sha256()
    with open(path, 'rb') as fh:
        for block in iter(lambda: fh.read(1024 * 1024), b''):
            h.update(block)
    return h.hexdigest()


def download(url, path):
    if not path.exists():
        urllib.request.urlretrieve(url, path)
    return {'url': url, 'file': str(path.relative_to(BASE)),
            'bytes': path.stat().st_size, 'sha256': sha256(path)}


def parse_series(path):
    meta, sample = {}, {}
    with gzip.open(path, 'rt', errors='replace') as fh:
        lines = fh.read().splitlines()
    for line in lines:
        if line.startswith(('!Series_', '!Sample_')):
            values = next(csv.reader([line], delimiter='\t'))
            target = sample if line.startswith('!Sample_') else meta
            target.setdefault(values[0].split('_', 1)[1], []).append(values[1:])
    start = next(i for i, s in enumerate(lines) if s == '!series_matrix_table_begin')
    end = next(i for i, s in enumerate(lines) if s == '!series_matrix_table_end')
    table = pd.read_csv(io.StringIO('\n'.join(lines[start + 1:end])), sep='\t',
                        index_col=0)
    table.index = table.index.astype(str)
    ids = sample['geo_accession'][0]
    records = []
    for i, gsm in enumerate(ids):
        record = {'sample_id': gsm, 'title': sample['title'][0][i],
                  'source_name': sample['source_name_ch1'][0][i]}
        for vals in sample.get('characteristics_ch1', []):
            key, _, value = vals[i].partition(': ')
            if key in record:
                raise ValueError(f'Duplicate characteristic {gsm}: {key}')
            record[key] = value
        record['description'] = ' | '.join(x[i] for x in sample.get('description', []))
        record['supplementary_file'] = ' | '.join(x[i] for x in sample.get('supplementary_file', []))
        records.append(record)
    assert ids == list(table.columns)
    return meta, pd.DataFrame(records), table.apply(pd.to_numeric, errors='raise')


def platform_mapping(platform):
    """Use only unambiguous nonempty symbols from the official GEO annotation."""
    filename = platform + '.annot.gz'
    local = SOURCE / filename
    url = f'https://ftp.ncbi.nlm.nih.gov/geo/platforms/{platform[:-3]}nnn/{platform}/annot/{filename}'
    path = local if local.exists() else DATA / filename
    if not path.exists():
        urllib.request.urlretrieve(url, path)
    mapping, omitted, source_rows = {}, 0, 0
    with gzip.open(path, 'rt', errors='replace') as fh:
        columns = None
        for line in fh:
            if line.startswith('!platform_table_begin'):
                columns = next(fh).rstrip('\n').split('\t')
                gene_index = next(i for i, s in enumerate(columns) if s.lower() == 'gene symbol')
                continue
            if line.startswith('!platform_table_end'):
                break
            if columns is None:
                continue
            values = line.rstrip('\n').split('\t')
            source_rows += 1
            if len(values) <= gene_index:
                omitted += 1
                continue
            symbols = sorted(set(s.strip() for s in re.split(r'///| // |;', values[gene_index])
                                 if s.strip() and s.strip() not in {'---', 'NA', 'na'}))
            if len(symbols) == 1:
                mapping[values[0].strip()] = symbols[0]
            else:
                omitted += 1
    return mapping, {'platform': platform, 'url': url, 'file': str(path),
                     'sha256': sha256(path), 'source_rows': source_rows,
                     'unambiguously_mapped_probes': len(mapping),
                     'unmapped_or_ambiguous_probes_excluded': omitted,
                     'mapping_rule': 'Only one unique nonempty official symbol; median across probes.'}


def gene_matrix(table, mapping):
    selected = table.loc[table.index.intersection(mapping)]
    symbols = pd.Index([mapping[p] for p in selected.index], name='gene')
    selected = selected.set_axis(symbols, axis=0)
    return selected.groupby(level=0).median().sort_index()


def main():
    cohorts, sample_tables = [], []
    for gse, platform in [('GSE41571', 'GPL570'), ('GSE21545', 'GPL570'),
                          ('GSE163154', 'GPL6104')]:
        filename = gse + '_series_matrix.txt.gz'
        url = f'https://ftp.ncbi.nlm.nih.gov/geo/series/{gse[:-3]}nnn/{gse}/matrix/{filename}'
        source = download(url, DATA / filename)
        series, samples, table = parse_series(DATA / filename)
        mapping, annotation = platform_mapping(platform)
        excluded = []
        supplemental_source = None
        if gse == 'GSE41571':
            # The series matrix is probe-wise median-relative expression, which
            # does not retain cross-gene abundance ordering. The depositor also
            # provides uncentered GCRMA-processed signal columns labelled "raw".
            # These are processed expression, NOT unprocessed CEL probe data.
            supp_name = 'GSE41571_matrix_NORMALIZED_RAW_ABSCALL.txt.gz'
            supp_url = f'https://ftp.ncbi.nlm.nih.gov/geo/series/GSE41nnn/GSE41571/suppl/{supp_name}'
            supplemental_source = download(supp_url, DATA / supp_name)
            supplemental = pd.read_csv(DATA / supp_name, sep='\t', index_col=0, dtype=str)
            for column in supplemental.columns:
                if column.endswith((' raw', ' normalized')):
                    # Commas are thousands separators; trailing '*' marks a
                    # floored normalized ratio. The latter is used only to
                    # cross-check GSM identity, never in the expression model.
                    supplemental[column] = pd.to_numeric(
                        supplemental[column].str.replace(',', '', regex=False).str.rstrip('*'), errors='raise')
            supplemental.index = supplemental.index.astype(str)
            samples['original_specimen_id'] = samples.supplementary_file.map(
                lambda s: re.search(r'_(CEA[rs]\d+)\.CEL', s).group(1))
            raw_columns, normalized_columns = {}, {}
            for row in samples.to_dict('records'):
                raw_columns[row['sample_id']] = supplemental[row['original_specimen_id'] + ' raw']
                normalized_columns[row['sample_id']] = supplemental[row['original_specimen_id'] + ' normalized']
            normalized = pd.DataFrame(normalized_columns).reindex(table.index)
            assert np.allclose(normalized.to_numpy(), table.to_numpy(), rtol=0, atol=0.000501), 'GSE41571 specimen-to-GSM mapping does not reproduce deposited normalized table within series rounding'
            table = pd.DataFrame(raw_columns).reindex(table.index).apply(pd.to_numeric, errors='raise')
            supplemental_source['columns_used'] = '11 columns ending in raw; identity mapped through per-GSM deposited CEL filenames'
            supplemental_source['identity_verification'] = 'All 601425 normalized values reproduce the GSM series matrix within 0.000501 (series rounds small ratios to 3 decimal places).'
            supplemental_source['rejected_columns'] = 'Series matrix and normalized columns: probe-wise median ratios are incompatible with abundance-rank transfer'
            supplemental_source['scale_note'] = 'Metadata calls RAW log-GCRMA values, but numerical values are positive uncentered intensities; preserve deposited scale and use per-sample ranks without assuming log2.'
            samples['label'] = samples['title'].map(
                lambda s: 1 if s.startswith('Ruptured Plaque,') else 0 if s.startswith('Stable Plaque,') else np.nan)
            assert samples.label.notna().all()
            # The publication describes 11 plaques, but does not supply donor identifiers.
            # Do not invent IDs or mistake within-group biological replicate numbers for donor IDs.
            samples['donor_id'] = ''
            samples['donor_status'] = 'one biological plaque per array; donor key not deposited'
            samples['endpoint_role'] = 'rupture-associated cross-bed and cell-compartment shift'
            samples['endpoint_name'] = 'histologically ruptured versus stable carotid plaque'
            samples['inference_unit'] = 'plaque sample; patient independence not directly auditable'
            role = 'secondary external rupture-associated transfer'
            caution = ('Laser-microdissected plaque macrophages, not whole tissue. Stable/ruptured '
                       'carotid plaques are biologically related to but not identical to AAA rupture. '
                       'Only 11 samples; publication describes biological plaques but no donor key is deposited.')
        elif gse == 'GSE21545':
            excluded = samples.loc[samples['sample type'] != 'carotid plaque', 'sample_id'].tolist()
            samples = samples.loc[samples['sample type'] == 'carotid plaque'].copy()
            samples['donor_id'] = gse + '_individual_' + samples['individual']
            assert not samples.donor_id.duplicated().any()
            samples['donor_status'] = 'explicit individual identifier; one plaque retained per individual'
            samples['label'] = samples['ischemic event'].map({'TRUE': 1, 'FALSE': 0})
            assert samples.label.notna().all()
            samples['followup_days'] = pd.to_numeric(samples['ischemic time (days)'], errors='raise')
            assert (samples.followup_days > 0).all()
            samples['endpoint_role'] = 'post-endarterectomy ischemic-event endpoint shift'
            samples['endpoint_name'] = 'future ischemic event with time-to-event/censoring'
            samples['inference_unit'] = 'individual with one carotid plaque sample'
            role = 'secondary prognostic endpoint-shift stress test'
            caution = ('Ischemic event TRUE/FALSE records follow-up events, NOT baseline symptoms or '
                       'plaque rupture. Event-free patients have variable censoring times: a binary AUROC '
                       'is descriptive and must not be represented as time-to-event validation. '
                       'Use time-to-event discrimination when estimating prognosis. Paired PBMC arrays '
                       'are excluded to avoid counting the same donor twice.')
        else:
            samples['label'] = samples['classification'].map({'IPH': 1, 'non-IPH': 0})
            assert samples.label.notna().all()
            samples['donor_id'] = ''
            samples['donor_status'] = '24 source patients with repeated lesion segments; GSM-to-donor crosswalk not deposited'
            samples['endpoint_role'] = 'intraplaque hemorrhage histology endpoint shift'
            samples['endpoint_name'] = 'IPH versus non-IPH carotid lesion segment'
            samples['inference_unit'] = 'lesion segment; repeated donor sampling confirmed, crosswalk unresolved'
            role = 'descriptive segment-level histology stress test only'
            caution = ('All source patients are symptomatic. IPH is not symptomatic status or rupture. '
                       'Original study PMC8236116 (DOI 10.1002/ctm2.458) confirms paired segments from '
                       '24 patients. Public GEO has 43 transcriptomic segments (16 non-IPH/27 IPH); '
                       'the published multimodal analysis retains 42 (16/26) after excluding one '
                       'IPH sample with failed mass spectrometry. GEO supplies no GSM-to-donor '
                       'crosswalk. Do not assume 43 independent patients, use sample-wise bootstrap '
                       'confidence intervals, or add 43 to pooled independent-patient totals.')
        samples['cohort'] = gse
        samples['label'] = samples.label.astype(int)
        matrix = gene_matrix(table.loc[:, samples.sample_id], mapping)
        assert not matrix.index.duplicated().any()
        assert np.isfinite(matrix.to_numpy()).all()
        assert list(matrix.columns) == samples.sample_id.tolist()
        expression_path = DATA / f'{gse}_expression.tsv.gz'
        sample_path = DATA / f'{gse}_samples.csv'
        matrix.to_csv(expression_path, sep='\t', compression={'method': 'gzip', 'mtime': 0}, float_format='%.10g')
        samples.to_csv(sample_path, index=False)
        meta_path = DATA / f'{gse}_series_metadata.json'
        meta_path.write_text(json.dumps(series, indent=2), encoding='utf-8')
        sample_tables.append(samples)
        cohorts.append({'accession': gse, 'platform': platform, 'role': role,
                        'geo_record': f'https://www.ncbi.nlm.nih.gov/geo/query/acc.cgi?acc={gse}',
                        'n_source_arrays': len(table.columns), 'n_included_samples': len(samples),
                        'n_negative': int((samples.label == 0).sum()),
                        'n_positive': int((samples.label == 1).sum()),
                        'n_genes': len(matrix), 'endpoint': samples.endpoint_name.iloc[0],
                        'source': source, 'annotation': annotation,
                        'supplemental_processed_source': supplemental_source,
                        'expression_file': str(expression_path.relative_to(BASE)),
                        'expression_sha256': sha256(expression_path),
                        'sample_file': str(sample_path.relative_to(BASE)),
                        'sample_sha256': sha256(sample_path),
                        'excluded_sample_ids': excluded,
                        'exclusion_reason': 'paired PBMC tissue; retain one plaque per explicit donor' if excluded else 'none',
                        'donor_key_available': bool((samples.donor_id != '').all()),
                        'caution': caution,
                        'series_pubmed_ids': series.get('pubmed_id', []),
                        'processed_data_policy': 'Depositor-normalized public expression; no extra between-sample correction; ranks computed by benchmark engine.'})
        if gse == 'GSE21545':
            cohorts[-1]['followup_audit'] = {
                'range_days': [int(samples.followup_days.min()), int(samples.followup_days.max())],
                'median_days': float(samples.followup_days.median()),
                'events_by_30_days': int(((samples.label == 1) & (samples.followup_days <= 30)).sum()),
                'landmark_30_days_eligible_donors': int((samples.followup_days > 30).sum()),
                'landmark_30_days_events': int(((samples.label == 1) & (samples.followup_days > 30)).sum()),
                'recommended_locked_sensitivity': '30-day landmark: retain followup_days >30 and subtract 30 from times; survival endpoint only; proposed from metadata before score inspection.'}
        print(gse, matrix.shape, samples.label.value_counts().to_dict(), flush=True)
    # Audit accession overlaps and exact GSM reuse against existing source cohorts.
    old_ids = set()
    old_accessions = ['GSE98278', 'GSE111782', 'GSE198600', 'GSE27034']
    for gse in old_accessions:
        with gzip.open(SOURCE / f'{gse}_series_matrix.txt.gz', 'rt') as fh:
            for line in fh:
                if line.startswith('!Sample_geo_accession'):
                    old_ids.update(next(csv.reader([line], delimiter='\t'))[1:])
                    break
    new_ids = [gsm for samples in sample_tables for gsm in samples.sample_id]
    assert len(new_ids) == len(set(new_ids)), 'GSM duplicated across extension cohorts'
    assert not (set(new_ids) & old_ids), 'GSM overlap with original cohorts'
    recruitment = []
    for gse in old_accessions + [c['accession'] for c in cohorts]:
        path = (SOURCE if gse in old_accessions else DATA) / f'{gse}_series_matrix.txt.gz'
        metadata = {}
        with gzip.open(path, 'rt', encoding='utf-8', errors='replace') as fh:
            for line in fh:
                if line.startswith('!series_matrix_table_begin'):
                    break
                if line.startswith('!Series_'):
                    values = next(csv.reader([line], delimiter='\t'))
                    metadata.setdefault(values[0][8:], []).extend(values[1:])
        recruitment.append({'accession': gse,
                            'contact_institute': metadata.get('contact_institute', []),
                            'contributors': metadata.get('contributor', []),
                            'pubmed_ids': metadata.get('pubmed_id', []),
                            'overall_design': metadata.get('overall_design', [])})
    publication_sources = []
    for pmid, pmcid in [('34185408', '8236116'), ('22371308', '3388132'), ('23122912', '3566542')]:
        p = DATA / f'PMC{pmcid}.xml'
        if p.exists():
            publication_sources.append({'pmid': pmid, 'pmcid': 'PMC' + pmcid,
                                        'public_url': f'https://pmc.ncbi.nlm.nih.gov/articles/PMC{pmcid}/',
                                        'retrieval_url': f'https://eutils.ncbi.nlm.nih.gov/entrez/eutils/efetch.fcgi?db=pmc&id={pmcid}&retmode=xml',
                                        'file': str(p.relative_to(BASE)), 'sha256': sha256(p)})
    audit = {'generated_at_utc': datetime.now(timezone.utc).isoformat(),
             'script_sha256': sha256(Path(__file__)),
             'selection_timing': 'New cohorts selected and metadata audited before any external model outcomes were inspected.',
             'data_scope': 'Public GEO processed expression and public metadata only.',
             'cohorts': cohorts,
             'publication_sources': publication_sources,
             'recruitment_and_publication_audit': recruitment,
             'publication_audit_findings': {
                 'GSE41571': 'Original Newcastle recruitment, PMID 23122912 / PMC3566542; 11 expression-array plaques, distinct from 14 additional qPCR validation plaques. No identifiable overlap with original cohorts.',
                 'GSE21545': 'BiKE Karolinska recruitment, PMID 22371308 / PMC3388132; 126 plaque donors and 97 paired PBMC. Mean follow-up 44 months, 25 future ischemic events (18 strokes, 7 MI).',
                 'GSE163154': 'Maastricht Human Plaque Study, PMID 34185408 / PMC8236116; 24 symptomatic patients, paired segments and subsequent exclusions/reclassification. Published multimodal n42 differs from deposited transcriptomic n43 as described above. This paper also reuses BiKE as external validation; that does not make BiKE and Maastricht the same cohort.'},
             'overlap_audit': {'original_accessions': old_accessions,
                               'exact_gsm_overlap': [],
                               'cross_study_donor_overlap': 'No shared source accession/GSM; recruiting organizations differ (Munich, Pittsburgh, Tulane, Mayo, Newcastle, Karolinska, Maastricht). No duplication identified from deposited study/publication provenance. Unavailable donor identities prevent definitive cross-study participant linkage.',
                               'GSE21545_internal_duplicates': '97 paired PBMC excluded; 126 unique plaque donor IDs retained.'},
             'considered_not_acquired': [{'accession': 'GSE28829',
                                         'reason': 'Early/advanced lesion stage differs from rupture; no need to expand beyond three preselected new cohorts.'}],
             'mapping_difference_from_original': 'Original run_analysis.py chooses the first symbol for multi-symbol probes. Extension excludes ambiguous multi-symbol probes. GPL570 retains 42,904/54,675 probe rows mapping to 20,848 genes; GPL6104 retains 20,502/22,185 probe rows mapping to 17,184 genes. Report this stricter extension preprocessing or apply a common rule in a sensitivity analysis.',
             'claims_not_supported': ['new clinical validation', 'universal panvascular predictor',
                                      '43 independent patients in GSE163154',
                                      'baseline symptomatic status inferred from GSE21545 follow-up event labels']}
    (BASE / 'cohort_audit.json').write_text(json.dumps(audit, indent=2), encoding='utf-8')
    summary_cols = ['accession', 'platform', 'role', 'n_source_arrays', 'n_included_samples',
                    'n_negative', 'n_positive', 'n_genes', 'endpoint', 'donor_key_available', 'caution']
    pd.DataFrame([{k: c[k] for k in summary_cols} for c in cohorts]).to_csv(BASE / 'cohort_audit.csv', index=False)
    pd.concat(sample_tables, ignore_index=True).to_csv(DATA / 'extension_samples_all.csv', index=False)
    (BASE / 'cohort_audit.txt').write_text('\n\n'.join(
        f"{c['accession']} | {c['role']} | n={c['n_included_samples']} ({c['n_positive']} positive / {c['n_negative']} negative)\n"
        f"Endpoint: {c['endpoint']}\n{c['caution']}\nGEO: {c['geo_record']}\n"
        f"Expression: {c['expression_file']}\nMetadata: {c['sample_file']}"
        for c in cohorts), encoding='utf-8')


if __name__ == '__main__':
    main()
```
