## Supplementary Data 1: analysis code, provenance and complete outputs for "Cross study transfer of vascular transcriptomic classifiers in seven public cohorts": download_public_inputs.py

```
"""Acquire the frozen public GEO inputs and reconstruct derived input tables.

Only official public NCBI URLs in public_input_snapshot.json are accessed.
Existing files are validated and never overwritten, including on mismatch.
Run --verify-existing for an offline, input-preserving check.
"""
from __future__ import annotations

import argparse
import csv
import gzip
import hashlib
import importlib.util
import io
import json
import os
from pathlib import Path
import shutil
import sys
import tarfile
import tempfile
import time
import urllib.parse
import urllib.request

HERE = Path(__file__).resolve().parent
ROOT = HERE.parent
SNAPSHOT_PATH = HERE / 'public_input_snapshot.json'
REPORT = HERE / 'public_input_verification.json'


def sha256(path):
    h = hashlib.sha256()
    with path.open('rb') as f:
        for chunk in iter(lambda: f.read(1024 * 1024), b''):
            h.update(chunk)
    return h.hexdigest()


def canonical_text_sha256(path):
    """Ignore gzip container metadata and CRLF versus LF text line endings."""
    data = gzip.open(path, 'rb').read() if path.suffix == '.gz' else path.read_bytes()
    return hashlib.sha256(data.replace(b'\r\n', b'\n')).hexdigest()


def local_path(relative):
    path = (ROOT / relative).resolve()
    path.relative_to(ROOT.resolve())
    return path


def check(path, item, derived=False):
    if not path.is_file():
        return {'path': item['path'], 'status': 'missing', 'valid': False}
    actual = sha256(path)
    result = {'path': item['path'], 'actual_sha256': actual, 'expected_sha256': item['sha256'],
              'bytes': path.stat().st_size, 'byte_exact': actual == item['sha256']}
    if derived:
        canonical = canonical_text_sha256(path)
        result['canonical_text_sha256'] = canonical
        result['canonical_text_equal'] = canonical == item['canonical_text_sha256']
        valid = result['canonical_text_equal']
    else:
        valid = result['byte_exact']
    result.update(valid=valid, status='verified' if valid else 'mismatch')
    return result


def validate_existing(snapshot):
    rows = [check(local_path(x['path']), x) for x in snapshot['raw_inputs']]
    rows += [check(local_path(x['path']), x, True) for x in snapshot['processed_inputs']]
    mismatches = [r['path'] for r in rows if r['status'] == 'mismatch']
    if mismatches:
        raise RuntimeError('Existing files differ from the locked public snapshot; none were overwritten. Inspect these paths: ' + ', '.join(mismatches))
    helper = HERE / 'cohort_extension.py'
    if not helper.exists() or sha256(helper) != snapshot['cohort_extension_script_sha256']:
        raise RuntimeError('cohort_extension.py does not match the acquisition snapshot. Restore the packaged version; inputs have not been overwritten.')
    return rows


def download(item):
    destination = local_path(item['path'])
    if destination.exists():
        return
    url = urllib.parse.urlparse(item['url'])
    if url.scheme != 'https' or url.hostname != 'ftp.ncbi.nlm.nih.gov':
        raise ValueError('Only the locked official NCBI HTTPS URLs are permitted')
    destination.parent.mkdir(parents=True, exist_ok=True)
    print(f'Download {item["path"]} ({item["bytes"]:,} bytes)', flush=True)
    fd, temporary_name = tempfile.mkstemp(prefix='.public-download-', dir=destination.parent)
    os.close(fd)
    temporary = Path(temporary_name)
    try:
        for attempt in range(3):
            try:
                request = urllib.request.Request(item['url'], headers={'User-Agent': 'VascBridge-reproducibility/1.0 (public GEO research)'})
                with urllib.request.urlopen(request, timeout=180) as response, temporary.open('wb') as out:
                    shutil.copyfileobj(response, out, length=1024 * 1024)
                break
            except Exception:
                if attempt == 2:
                    raise
                time.sleep(2 * (attempt + 1))
        if sha256(temporary) != item['sha256']:
            raise RuntimeError(f'Download SHA-256 mismatch for {item["path"]}; remote metadata/content may have changed. No existing input was replaced.')
        # Exclusive final creation prevents accidental replacement if another
        # process creates the destination while this download is running.
        with destination.open('xb') as out, temporary.open('rb') as inp:
            shutil.copyfileobj(inp, out, length=1024 * 1024)
    finally:
        if temporary.exists():
            temporary.unlink()


def build_fpkm(snapshot):
    expected = next(x for x in snapshot['processed_inputs'] if x['path'] == 'GSE198600_fpkm.tsv')
    output = local_path(expected['path'])
    if output.exists():
        return
    archive = ROOT / 'GSE198600_RAW.tar'
    print('Reconstruct GSE198600 FPKM table from public supplementary archive', flush=True)
    rows, gene_order, samples = {}, [], []
    with tarfile.open(archive) as tar:
        for member in sorted(tar.getmembers(), key=lambda x: x.name):
            if not member.isfile():
                continue
            sample = Path(member.name).name.split('_')[0]
            if not sample.startswith('GSM') or sample in samples:
                raise ValueError(f'Unexpected or duplicate sample archive member: {member.name}')
            samples.append(sample)
            # Stream members without extracting paths from the archive.
            with gzip.GzipFile(fileobj=tar.extractfile(member)) as gz:
                reader = csv.DictReader(io.TextIOWrapper(gz, encoding='utf-8', errors='replace'), delimiter='\t')
                for record in reader:
                    gene = record.get('gene_id', '')
                    if gene not in rows:
                        rows[gene] = {}
                        gene_order.append(gene)
                    rows[gene][sample] = record.get('FPKM', '')
    fd, name = tempfile.mkstemp(prefix='.GSE198600_fpkm-', suffix='.tsv', dir=ROOT)
    os.close(fd)
    temporary = Path(name)
    try:
        with temporary.open('w', newline='', encoding='utf-8') as f:
            writer = csv.writer(f, delimiter='\t')
            writer.writerow(['gene_id'] + samples)
            for gene in gene_order:
                writer.writerow([gene] + [rows[gene].get(sample, '') for sample in samples])
        verified = check(temporary, expected, True)
        if not verified['valid']:
            raise RuntimeError('Reconstructed FPKM table differs from locked canonical content; no input was replaced')
        with output.open('xb') as out, temporary.open('rb') as inp:
            shutil.copyfileobj(inp, out)
    finally:
        if temporary.exists():
            temporary.unlink()


def build_extensions(snapshot):
    expected = [x for x in snapshot['processed_inputs'] if x['path'].startswith('bmc_revision/')]
    if all(local_path(x['path']).exists() for x in expected):
        return
    print('Reconstruct three external cohorts in an isolated staging directory', flush=True)
    # The frozen public-data transformation helper is run unchanged. Redirect
    # its output globals into staging, so existing derived files stay intact.
    with tempfile.TemporaryDirectory(prefix='.public-input-stage-', dir=HERE) as tmp:
        stage = Path(tmp).resolve()
        stage.relative_to(HERE.resolve())  # bounded cleanup target
        stage_data = stage / 'data'
        stage_data.mkdir()
        for item in snapshot['raw_inputs']:
            if item['path'].startswith('bmc_revision/data/'):
                src = local_path(item['path'])
                dst = stage_data / src.name
                try:
                    os.link(src, dst)
                except OSError:
                    shutil.copy2(src, dst)
        spec = importlib.util.spec_from_file_location('public_cohort_extension_rebuild', HERE / 'cohort_extension.py')
        module = importlib.util.module_from_spec(spec)
        spec.loader.exec_module(module)
        module.BASE, module.DATA, module.SOURCE = stage, stage_data, ROOT
        module.main()
        for item in expected:
            generated = stage_data / Path(item['path']).name
            if not check(generated, item, True)['valid']:
                raise RuntimeError(f'Reconstructed table differs from locked canonical content: {item["path"]}; existing input files unchanged')
        # Validate all generated inputs first, then promote only absent files.
        for item in expected:
            destination = local_path(item['path'])
            if not destination.exists():
                with destination.open('xb') as out, (stage_data / destination.name).open('rb') as inp:
                    shutil.copyfileobj(inp, out)
        # Provenance files are useful but are never allowed to overwrite the
        # reviewed original audit. Acquisition verification is separately saved.
        for name in ['cohort_audit.json', 'cohort_audit.csv', 'cohort_audit.txt']:
            destination = HERE / name
            if not destination.exists():
                if name.endswith('.json'):
                    audit = json.loads((stage / name).read_text(encoding='utf-8'))
                    for cohort in audit['cohorts']:
                        annotation_name = Path(cohort['annotation']['file']).name
                        cohort['annotation']['file'] = '../' + annotation_name if (ROOT / annotation_name).exists() else 'data/' + annotation_name
                    text = json.dumps(audit, indent=2)
                    with destination.open('x', encoding='utf-8') as out:
                        out.write(text)
                else:
                    with destination.open('xb') as out, (stage / name).open('rb') as inp:
                        shutil.copyfileobj(inp, out)


def main():
    parser = argparse.ArgumentParser(description=__doc__)
    parser.add_argument('--verify-existing', action='store_true', help='Offline check; no network, no reconstruction, no input replacement')
    args = parser.parse_args()
    snapshot = json.loads(SNAPSHOT_PATH.read_text(encoding='utf-8'))
    rows = validate_existing(snapshot)
    if not args.verify_existing:
        for item in snapshot['raw_inputs']:
            download(item)
        build_fpkm(snapshot)
        build_extensions(snapshot)
        rows = validate_existing(snapshot)
    missing = [r['path'] for r in rows if not r['valid']]
    report = {'snapshot': SNAPSHOT_PATH.name, 'snapshot_sha256': sha256(SNAPSHOT_PATH),
              'mode': 'offline verification' if args.verify_existing else 'download missing public inputs and reconstruct missing derived inputs',
              'all_verified': not missing, 'raw_input_count': len(snapshot['raw_inputs']),
              'derived_input_count': len(snapshot['processed_inputs']), 'files': rows,
              'compressed_output_policy': 'Public downloaded files require exact byte hashes. Derived inputs allow gzip-header/newline differences only if canonical decompressed text SHA-256 is identical.',
              'existing_files_overwritten': False}
    REPORT.write_text(json.dumps(report, indent=2), encoding='utf-8')
    if missing:
        print('Missing/unverified inputs: ' + ', '.join(missing), file=sys.stderr)
        return 1
    print(f'PASS: {len(rows)} public/derived inputs verified; no existing input overwritten. Report: {REPORT.name}', flush=True)
    return 0


if __name__ == '__main__':
    raise SystemExit(main())
```
