## Supplementary Data 1: analysis code, provenance and complete outputs for "Cross study transfer of vascular transcriptomic classifiers in seven public cohorts": editorial_result_review.py

```
"""Independent review of saved result outputs; does not refit or alter results."""
import pathlib,json
import numpy as np,pandas as pd
from sklearn.metrics import roc_auc_score,average_precision_score,brier_score_loss
here=pathlib.Path(__file__).parent;out=here/'results'
m=pd.read_csv(out/'metrics.csv',float_precision='round_trip')
p=pd.read_csv(out/'predictions.csv',float_precision='round_trip')
checks=[]
for _,r in m[m.cohort!='GSE98278'].iterrows():
 q=p[(p.variant==r.variant)&(p.model==r.model)&(p.cohort==r.cohort)&(p.prediction_type=='frozen_external')].copy()
 if r.cohort=='GSE21545' and r.subset=='30_day_landmark':q=q[q.followup_days>30]
 assert len(q)==r.n and q.sample_id.is_unique
 y=q.label.to_numpy(int);s=q.score.to_numpy(float)
 assert np.isfinite(s).all()
 if r.cohort!='GSE21545':
  auc=roc_auc_score(y,s);ap=average_precision_score(y,s)
  assert abs(auc-r.auroc)<1e-12 and abs(ap-r.auprc)<1e-12
  if np.isfinite(r.brier):assert abs(brier_score_loss(y,s)-r.brier)<1e-12
  checks.append({'variant':r.variant,'model':r.model,'cohort':r.cohort,'n':len(q),'auc':auc,'all_metrics_match':True})
 else:
  t=q.followup_days.to_numpy(float)
  comparable=[(i,j) for i in range(len(t)) for j in range(i+1,len(t)) if (y[i] and t[i]<t[j]) or (y[j] and t[j]<t[i])]
  num=0
  for i,j in comparable:
   early,late=(i,j) if t[i]<t[j] else (j,i)
   num+=(s[early]>s[late])+0.5*(s[early]==s[late])
  c=num/len(comparable);assert abs(c-r.harrell_c)<1e-12
  ec_ties=sum(t[i]==t[j] and y[i]!=y[j] for i in range(len(t)) for j in range(i+1,len(t)))
  checks.append({'variant':r.variant,'model':r.model,'cohort':r.cohort,'subset':r.subset,'n':len(q),'c_index':c,'comparable_pairs':len(comparable),'tied_event_censor_pairs_excluded':ec_ties,'all_metrics_match':True})
q=p[(p.variant=='source_rank')&(p.model=='L1_logistic')&(p.cohort=='GSE41571')&(p.prediction_type=='frozen_external')]
perfect={'n':len(q),'auroc':roc_auc_score(q.label,q.score),'score_min':q.score.min(),'score_max':q.score.max(),'at_05_sensitivity':float((q[q.label==1].score>=.5).mean()),'at_05_specificity':float((q[q.label==0].score<.5).mean()),'case_min':float(q[q.label==1].score.min()),'control_max':float(q[q.label==0].score.max())}
rep={'external_metric_checks_passed':True,'n_rows_checked':len(checks),'checks':checks,'GSE41571_source_rank_L1':perfect,'source_run_summary_rows':json.loads((out/'run_summary.json').read_text())['n_metrics'],'reporting_audit_rows':len(m)}
(here/'editorial_result_audit.json').write_text(json.dumps(rep,indent=2,default=lambda x:x.item()),encoding='utf-8')
print(json.dumps({'external_metric_checks_passed':True,'n_rows_checked':len(checks),'GSE41571_source_rank_L1':perfect,'tied_event_censor_pairs':sorted(set(x.get('tied_event_censor_pairs_excluded',0) for x in checks))},indent=2,default=lambda x:x.item()))
```
