## Supplementary Data 1: analysis code, provenance and complete outputs for "Cross study transfer of vascular transcriptomic classifiers in seven public cohorts": make_revision_figures.py

```
"""Publication figures drawn exclusively from saved revision outputs."""
from pathlib import Path
import json
import numpy as np
import pandas as pd
import matplotlib
matplotlib.use('Agg')
import matplotlib.pyplot as plt
from matplotlib.patches import Rectangle

BASE=Path(__file__).resolve().parent
R=BASE/'results'; F=BASE/'figures'; F.mkdir(exist_ok=True)
M=pd.read_csv(R/'metrics.csv',float_precision='round_trip')
P=pd.read_csv(R/'predictions.csv',float_precision='round_trip')
# Draw at the actual 6.4-inch manuscript width: no downstream font shrinkage.
plt.rcParams.update({'font.family':'DejaVu Sans','font.size':8.5,'axes.spines.top':False,
 'axes.spines.right':False,'axes.titleweight':'bold','axes.titlesize':9.7,
 'axes.labelsize':8.5,'xtick.labelsize':8.5,'ytick.labelsize':8.5,
 'legend.fontsize':8.2,'pdf.fonttype':42,'ps.fonttype':42,
 'savefig.facecolor':'white','axes.linewidth':.7})
COL=['#16697A','#E58F20','#85579B','#3F8D63','#8C959D']
MODELS=['L1_logistic','elastic_net','linear_SVM','random_forest','dummy_prior']
NAMES={'L1_logistic':'L1 logistic','elastic_net':'Elastic net','linear_SVM':'Linear SVM',
 'random_forest':'Random forest','dummy_prior':'Prior only'}
def metric(model,cohort,variant='source_rank',subset='all'):
    s=M[(M.model==model)&(M.cohort==cohort)&(M.variant==variant)&(M.subset==subset)]
    assert len(s)==1,(model,cohort,variant,subset)
    return s.iloc[0]
def save(fig,name):
    # Preserve the physical canvas width, so 8.5-pt text remains 8.5 pt in Word.
    fig.savefig(F/(name+'.pdf'))
    fig.savefig(F/(name+'.png'),dpi=300)
    plt.close(fig)
def panel(ax,label,title):
    ax.set_title(label+'  '+title,loc='left',pad=9)
def heat(ax,values,rows,cols,vmin=0,vmax=1):
    im=ax.imshow(values,cmap='RdBu',vmin=vmin,vmax=vmax,aspect='auto')
    ax.set_xticks(range(len(cols)),cols); ax.set_yticks(range(len(rows)),rows)
    for i in range(len(rows)):
        for j in range(len(cols)):
            v=values[i,j]
            ax.text(j,i,f'{v:.3f}',ha='center',va='center',fontsize=8.8,color='white' if v<.2 or v>.8 else '#111111')
    return im

# Figure 1: study design, units and distinct endpoints.
fig=plt.figure(figsize=(6.4,6.0))
a=fig.add_axes([.045,.73,.91,.20]); b=fig.add_axes([.335,.195,.60,.43])
a.axis('off'); panel(a,'A','Source development and frozen evaluation')
boxes=[(.00,'GSE98278\n48 AAA samples\n31 stable / 17 ruptured'),(.35,'Source-only selection\nOuter CV: 4 folds x 3\nInner tuning: 3 folds\n4 classifiers + prior'),(.70,'Freeze each model\nSeparate external tests\nNo external tuning\nNo endpoint pooling')]
for xx,txt in boxes:
    a.add_patch(Rectangle((xx,.12),.30,.77,transform=a.transAxes,facecolor='#EEF4F6',edgecolor='#16697A',lw=.9))
    a.text(xx+.15,.505,txt,ha='center',va='center',transform=a.transAxes,fontsize=8.5,linespacing=1.35)
for x1,x2 in [(.305,.345),(.655,.695)]:
    a.annotate('',xy=(x2,.505),xytext=(x1,.505),xycoords='axes fraction',arrowprops={'arrowstyle':'->','color':'#16697A','lw':1.2})
labels=['Carotid symptoms\nGSE111782','Recent carotid event\nGSE198600','Macrophage rupture\nGSE41571','Future ischaemic events\nGSE21545','IPH / non-IPH segments\nGSE163154','PAD processing control\nGSE27034']
negative=np.array([9,5,6,101,16,18]); positive=np.array([9,6,5,25,27,19]); yy=np.arange(6)
b.barh(yy,negative,color='#BCD2DC',label='Reference / censored')
b.barh(yy,positive,left=negative,color='#16697A',label='Case / event / IPH')
for i,n in enumerate(negative+positive):b.text(n+2,i,str(n),va='center',fontsize=8.5)
b.set_yticks(yy,labels); b.invert_yaxis(); b.set_xlim(0,151); b.set_xlabel('Deposited observations included')
b.legend(frameon=False,loc='upper center',bbox_to_anchor=(.40,-.21),ncol=2,columnspacing=1.4)
panel(b,'B','Different endpoints and observation units')
fig.text(.045,.026,'Counts are observations, not a pooled patient population. IPH segments come from\n24 source patients. PAD has incompatible preprocessing.',fontsize=8,linespacing=1.4)
save(fig,'Figure1_design')

# Figure 2: nested source and endpoint-specific external comparison.
fig,(a,b)=plt.subplots(2,1,figsize=(6.4,6.3),gridspec_kw={'height_ratios':[.85,1.15]})
fig.subplots_adjust(left=.20,right=.90,bottom=.17,top=.94,hspace=.62)
for i,model in enumerate(MODELS):
    r=metric(model,'GSE98278')
    a.errorbar(r.auroc,i,xerr=[[r.auroc-r.auroc_ci_low],[r.auroc_ci_high-r.auroc]],fmt='o',color=COL[i],capsize=3)
a.set_yticks(range(5),[NAMES[x] for x in MODELS]); a.invert_yaxis(); a.set_xlim(0,1.03); a.axvline(.5,color='#999999',ls='--',lw=1)
a.set_xlabel('AUROC'); panel(a,'A','Nested source OOF scores')
cs=['GSE111782','GSE198600','GSE41571','GSE163154']
v=np.array([[metric(model,c).auroc for c in cs] for model in MODELS])
im=heat(b,v,[NAMES[x] for x in MODELS],['Symptoms\nn=18','Recent event\nn=11','Macrophage\nrupture: n=11','IPH segments\nn=43*'])
panel(b,'B','Frozen models across external endpoints'); fig.colorbar(im,ax=b,fraction=.035,pad=.03,label='AUROC')
fig.text(.045,.025,'Source bars: 95% bootstrap intervals conditional on OOF scores.\n*IPH estimates are descriptive; the donor-to-segment mapping is unresolved.',fontsize=8,linespacing=1.4)
save(fig,'Figure2_model_comparison')

# Figure 3: joint pipeline sensitivity and active feature coverage.
fig,(a,b)=plt.subplots(2,1,figsize=(6.4,6.15),gridspec_kw={'height_ratios':[.78,1.1]})
fig.subplots_adjust(left=.24,right=.95,bottom=.17,top=.94,hspace=.73)
variants=['source_rank','common_rank','source_log']; cs=['GSE98278','GSE111782','GSE198600','GSE41571']
v=np.array([[metric('L1_logistic',c,variant).auroc for c in cs] for variant in variants])
im=heat(a,v,['Source ranks','Common ranks','Scale diagnostic'],['AAA\nn=48','Carotid array\nn=18','Carotid RNA\nn=11','Macrophages\nn=11'])
panel(a,'A','Separately source-tuned L1 pipelines')
cov=pd.read_csv(R/'active_feature_missingness.csv')
cohorts=['GSE111782','GSE198600','GSE41571','GSE163154','GSE21545']
for i,model in enumerate(MODELS[:4]):
    z=cov[(cov.variant=='source_rank')&(cov.model==model)].set_index('cohort')
    b.plot(range(len(cohorts)),[100*z.loc[c,'absolute_weight_missing_fraction'] for c in cohorts],marker='o',label=NAMES[model],color=COL[i])
b.set_xticks(range(5),['Carotid\narray','Carotid\nRNA','Macrophages','IPH\nsegments','Follow-up']); b.set_ylim(-1,40)
b.set_ylabel('Missing absolute weight\nor importance (%)'); b.legend(frameon=False,ncol=2,loc='upper right',columnspacing=1.1); panel(b,'B','Missing genes across fitted models')
fig.text(.045,.025,'These joint pipeline comparisons do not isolate a causal mechanism.\nThe primary L1 model has three active genes, all measured in the external cohorts.',fontsize=8,linespacing=1.4)
save(fig,'Figure3_pipeline_sensitivity')

# Figure 4: survival discrimination with censoring retained.
fig,axs=plt.subplots(1,2,figsize=(6.4,3.6),sharey=True)
fig.subplots_adjust(left=.18,right=.97,bottom=.28,top=.78,wspace=.22)
for ax,subset,title in zip(axs,['all','30_day_landmark'],['A  All follow-up\n126 donors, 25 events','B  30-day landmark\n114 donors, 13 events']):
    for i,model in enumerate(MODELS[:4]):
        r=metric(model,'GSE21545',subset=subset)
        ax.errorbar(r.harrell_c,i,xerr=[[r.harrell_c-r.c_ci_low],[r.c_ci_high-r.harrell_c]],fmt='o',color=COL[i],capsize=3)
    ax.set_title(title,loc='left',pad=10); ax.set_yticks(range(4),[NAMES[x] for x in MODELS[:4]]); ax.set_xlim(.15,.85); ax.set_xticks([.2,.4,.6,.8]); ax.axvline(.5,color='#999999',ls='--',lw=.9); ax.set_xlabel('Harrell concordance index')
axs[0].invert_yaxis(); fig.text(.045,.035,'Bars: 95% donor-bootstrap intervals conditional on frozen scores.\nCensoring is retained; the two analyses use overlapping donors.',fontsize=8,linespacing=1.4)
save(fig,'Figure4_survival')

# Figure 5: diameter strata and the different questions asked by subgroup tests.
fig=plt.figure(figsize=(6.4,5.65))
a=fig.add_axes([.15,.61,.80,.27]); b=fig.add_axes([.24,.27,.71,.22])
cats=['intermediate','large','unknown']; ctrl=[15,7,9]; case=[0,9,8]; x=np.arange(3)
a.bar(x-.17,ctrl,.34,label='Elective stable',color='#BCD2DC'); a.bar(x+.17,case,.34,label='Ruptured',color='#16697A')
a.set_xticks(x,['Intermediate','Large','Unknown']); a.set_ylim(0,18); a.set_ylabel('Source tissue samples'); a.legend(frameon=False,loc='upper right'); panel(a,'A','Size categories and rupture status')
full=pd.read_csv(R/'full_source_oof_diameter_sensitivity.csv')
for i,cat in enumerate(['large','unknown']):
    r=M[(M.variant=='source_rank')&(M.model=='L1_logistic')&(M.cohort=='GSE98278')&(M.subset=='diameter_'+cat)].iloc[0]
    f=full[full.subset=='full_source_oof_within_'+cat].iloc[0]
    b.errorbar(f.auroc,i-.10,xerr=[[f.auroc-f.auroc_ci_low],[f.auroc_ci_high-f.auroc]],fmt='o',color=COL[0],capsize=3,label='Full-source OOF, within category' if i==0 else '')
    b.errorbar(r.auroc,i+.10,xerr=[[r.auroc-r.auroc_ci_low],[r.auroc_ci_high-r.auroc]],fmt='s',color=COL[1],capsize=3,label='Retrained within category' if i==0 else '')
b.set_yticks([0,1],['Large: n=16','Unknown: n=17']); b.invert_yaxis(); b.set_xlim(0,1.05); b.axvline(.5,color='#999999',ls='--',lw=.9); b.set_xlabel('AUROC'); b.legend(frameon=False,loc='upper center',bbox_to_anchor=(.40,-.35),ncol=2,columnspacing=1.1); panel(b,'B','Subgroup evaluation and retraining differ')
fig.text(.045,.025,'Restricted retraining changes training size and composition. This contrast does\nnot identify the causal effect of aneurysm diameter.',fontsize=8,linespacing=1.4)
save(fig,'Figure5_diameter')

# Figure 6: correctly matched fixed-baseline permutation control and tiny-cohort scores.
fig,(a,b)=plt.subplots(1,2,figsize=(6.4,3.65))
fig.subplots_adjust(left=.10,right=.97,bottom=.29,top=.79,wspace=.45)
null=pd.read_csv(R/'permutation_null.csv'); ps=json.loads((R/'permutation_summary.json').read_text(encoding='utf-8'))
a.hist(null.auroc,bins=np.linspace(0,1,21),color='#BCD2DC',edgecolor='white')
a.axvline(ps['observed_auroc'],color='#16697A',lw=2,label=f"Observed {ps['observed_auroc']:.3f}")
a.set_xlim(0,1); a.set_ylim(0,38); a.set_xlabel('Fixed baseline AUROC'); a.set_ylabel('Permutations'); a.legend(frameon=False,fontsize=8.2); panel(a,'A','Fixed-baseline control\n200 label permutations')
d=P[(P.variant=='source_rank')&(P.model=='L1_logistic')&(P.cohort=='GSE41571')]
for label,col in [(0,'#BCD2DC'),(1,'#16697A')]:
    vals=d[d.label==label].score.to_numpy(); xx=label+np.linspace(-.09,.09,len(vals))
    b.scatter(xx,vals,s=35,color=col,edgecolor='#333333',linewidth=.5)
b.set_xticks([0,1],['Stable\nn=6','Ruptured\nn=5']); b.set_xlim(-.45,1.45); b.set_ylim(0,1); b.set_ylabel('Frozen L1 model score'); panel(b,'B','Macrophage score ranking\n11 samples only')
fig.text(.045,.028,'Permutation P = 1/201 concerns the fixed baseline, not the tuned models.\nPanel B does not establish calibration or population-level certainty.',fontsize=8,linespacing=1.4)
save(fig,'Figure6_uncertainty')
print('Saved six PDF and PNG figures to',F)
```
