## Supplementary Data 1: analysis code, provenance and complete outputs for "Cross study transfer of vascular transcriptomic classifiers in seven public cohorts": README_REPRODUCE.txt

VascBridge-Instability: reproducible public-data revision
======================================================

This package contains analysis code, protocol/amendment records, and small
results. It does not redistribute the large expression input files. All input
expression and sample metadata are publicly available from NCBI GEO. No private
clinical data, institutional account, or paid dataset is required.

LAYOUT
------
Preserve the bmc_revision directory name. Its parent is the analysis root:

  analysis_root/
    bmc_revision/
      download_public_inputs.py
      public_input_snapshot.json
      cohort_extension.py
      benchmark_revision.py
      audit_results.py
      requirements-lock.txt
      requirements-reporting.txt
      data/       (downloaded/reconstructed external inputs)
      results/    (analysis outputs)
    GSE98278_series_matrix.txt.gz  (downloaded source inputs appear here)
    ...

Commands below are run from analysis_root. Script paths are resolved relative
to their own location, so another working directory is also supported when
absolute script paths are supplied. The acquisition step downloads about
152 MB of compressed public inputs; allow at least 1 GB free disk and 2 GB RAM.

ENVIRONMENT
-----------
The completed analysis used Python 3.11 on Windows. The exact scientific
package versions are recorded in requirements-lock.txt and
results/environment.json. Create an isolated Python environment, then run:

  python -m pip install -r bmc_revision/requirements-lock.txt

For example, environment activation is platform-specific:
  Windows: python -m venv .venv
           .venv\Scripts\activate
  Linux/macOS: python3 -m venv .venv
               source .venv/bin/activate

ACQUIRE AND VERIFY PUBLIC INPUTS
-------------------------------
  python bmc_revision/download_public_inputs.py

This downloads only missing files from the official HTTPS NCBI FTP service.
It obtains four original series matrices, three original platform annotations,
the GSE198600 public RAW supplementary archive, three extension matrices,
GPL6104 annotation, and the GSE41571 processed-signal supplement. It reconstructs
GSE198600_fpkm.tsv by sample accession, and executes the frozen
cohort_extension.py in an isolated staging directory to construct three
gene-by-sample external matrices plus their explicit endpoint metadata.

public_input_snapshot.json locks URLs and expected hashes. Every pre-existing
input is checked before any downloads. A mismatch stops the command without
overwriting the mismatched file. If GEO changes a compressed archive or
annotation, investigate and document the change before using a different
snapshot; do not silently accept it as the original analysis.

Downloaded raw/processed-source files require exact byte hashes. Reconstructed
tables also carry canonical SHA-256 hashes after decompression and CRLF-to-LF
normalization. Thus a different gzip header or operating-system line ending
does not falsely imply altered expression values. Numeric and textual content
must still match exactly. The actual hashes are recorded in
public_input_verification.json. Existing inputs and the reviewed cohort audit
are not overwritten.

To verify an already populated workspace without downloading or rebuilding:
  python bmc_revision/download_public_inputs.py --verify-existing

RUN THE ANALYSIS
----------------
  python bmc_revision/benchmark_revision.py
  python bmc_revision/audit_results.py

The first command writes results/ and refits the declared analysis. It uses
four outer folds repeated three times and three inner folds, with all
imputation, variance selection, supervised selection and scaling fitted only
on training data. Models and finite hyperparameter grids are in
analysis_protocol.json. The 200 whole-pipeline permutations assess a separate
fixed L1 baseline (k=100, C=0.25), not the tuned-model selection procedure.

The second command preserves pre-reporting metrics, suppresses degenerate
bootstrap intervals, evaluates full-source OOF scores within available size
categories, audits active-feature missingness, and creates the reproducibility
manifest. It does not refit or optimize any model. Run in a fresh copy when
comparing to the archived results, because the benchmark intentionally writes
its results directory. The reporting audit reads its raw preserved metrics
when rerun; do not combine raw metrics from another code/data version.

The archived run_summary.json also inventories two public files found in the
original workspace (GSE100927 series matrix and GPL17077 annotation) that the
revision engine does not read. They are not required and the downloader does
not fetch them. A clean run's input-file inventory therefore excludes these
unused files. Historical hashes describe the original run; operating-system
details, acquisition timestamps, and generated-container hashes can differ
without a numerical change. The supplementary archive's own MANIFEST_SHA256.json
describes the exact distributed bytes.

Useful output files:
  results/metrics.csv                    final 61 metric rows
  results/predictions.csv                sample-level model predictions
  results/nested_folds.csv               split membership and inner choices
  results/permutation_summary.json       200-permutation summary and scope
  results/permutation_sample_predictions.csv.gz
  results/missing_gene_audit.csv         selected-gene coverage
  results/active_feature_missingness.csv nonzero-weight coverage
  results/reproducibility_manifest.json  source and output hashes

When reading numeric CSV outputs using pandas, specify
float_precision='round_trip'. Near-equal random-forest scores can otherwise
round into ties during CSV parsing and slightly alter an AUROC recomputation.
Uncompressed file hashes may differ with platform line endings; compare
canonical input content and numerical results, not gzip container timestamps.

FIGURES AND INDEPENDENT OUTPUT CHECKS
------------------------------------
Figure generation has an additional plotting dependency kept separate from
the scientific-computation lock:

  python -m pip install -r bmc_revision/requirements-reporting.txt
  python bmc_revision/make_revision_figures.py

This reads only saved results and creates six PDF/PNG figures under
bmc_revision/figures/. It does not access raw inputs or refit models.
The archived small-output audit can also be rerun without downloading inputs:

  python bmc_revision/editorial_result_review.py

Frozen .joblib models are included for inspection in the matching environment.
They contain the custom TrainOnlySelect class serialized from the executable
analysis module. To load them in an interactive Python session from the root:

  import __main__, joblib
  from bmc_revision.benchmark_revision import TrainOnlySelect
  __main__.TrainOnlySelect = TrainOnlySelect
  model = joblib.load('bmc_revision/results/frozen_source_rank_L1_logistic.joblib')

Inputs must still undergo the saved source-vocabulary transformation in
benchmark_revision.py; a model file alone is not a clinical prediction service.
Only load model files from a trusted archive. The preferred full reproduction
is the explicit acquisition and refitting workflow above.

PACKAGE CONTENTS
----------------
Additional file 1 preserves the bmc_revision/ directory. It includes all small
numeric outputs and frozen models, public-cohort metadata audits, executable
acquisition and analysis scripts, and computational/reporting checks. Large
expression inputs and manuscript assembly scripts are excluded. Audit copies
may normalize local annotation paths to relative references and remove
editorial work-status notes; scientific results and computation scripts are
included unchanged. PACKAGE_NOTES.json records those packaging-only edits.
These internal checks are not external peer review.

SCIENTIFIC LIMITS THAT MUST TRAVEL WITH THE RESULTS
--------------------------------------------------
This is a post-review revision, not a preregistration or clinical validation.
GSE111782, GSE198600 and GSE27034 were previously inspected external cohorts.
No external labels are used for model fitting or hyperparameter selection.

GSE41571 uses the depositor's uncentered processed-signal columns labelled
'raw', not its per-probe-median ratios and not raw CEL probe processing. Its
11 plaque samples are too small to establish reliable clinical prediction;
perfect observed discrimination gives a degenerate empirical bootstrap.

GSE27034 contains per-probe-centered values, which do not preserve cross-gene
abundance ranks. It is a preprocessing-incompatible stress control and cannot
validate biological cross-bed transfer.

GSE163154 contains 43 segments from a study of 24 patients with repeated
sampling. The public sample-to-donor crosswalk is unavailable. Only descriptive
segment AUROC is reported; no independent-patient confidence interval is used.

GSE21545 contributes 126 unique plaque donors with future events/censoring.
Harrell C-index is evaluated using time and event indicators, including a
30-day landmark. Censored individuals are not treated as ordinary binary
event-free controls. The paired PBMC arrays are excluded.

GSE98278 lacks a deposited donor identifier. Cross-validation and bootstrap
uncertainty are sample-level, with independence not directly verifiable.
The diameter categories are incomplete and unevenly distributed. Separately
refitting tiny size strata is not equivalent to evaluating source-wide OOF
scores within those strata; neither isolates a causal diameter effect.

DECLARATION OF GENERATIVE AI AND AI-ASSISTED TECHNOLOGIES
Codex assisted with LaTeX assembly, prose editing, and consistency checks. The authors reviewed and revised all AI-assisted text and take responsibility for the submitted manuscript.
