## Supplementary Data 1: analysis code, provenance and complete outputs for "Cross study transfer of vascular transcriptomic classifiers in seven public cohorts": README_PRIVACY_RELEASE_20261029.txt

START HERE: medRxiv Supplementary Data 1 privacy/file-format correction
Manuscript MEDRXIV/2026/366574
Cross study transfer of vascular transcriptomic classifiers in seven public cohorts
Release date: 29 October 2026

This is a distribution-only correction to the previously uploaded archive.
The manuscript, scientific calculations, predictions, metric values, source
code, seeds, folds and interpretation have not been changed or refitted.
This file and PRIVACY_RELEASE_MANIFEST_20261029.json describe THIS release.

WHAT WAS REMOVED
Five redundant metadata CSV files carrying unused exact-age characteristics
are omitted, including two copies in results/ that duplicate the input ages:
  bmc_revision/data/GSE21545_samples.csv
  bmc_revision/data/GSE41571_samples.csv
  bmc_revision/data/extension_samples_all.csv
  bmc_revision/results/GSE21545_sample_manifest.csv
  bmc_revision/results/GSE41571_sample_manifest.csv
Seven frozen .joblib model files are omitted because this release contains
text-screenable materials only. They were saved outputs, not prerequisites
for recomputing the reported metrics from the saved predictions. Do not
interpret the absence of model binaries as a change in the model methods.

The only nested gzip payload, permutation_sample_predictions.csv.gz, has
been expanded into permutation_sample_predictions.csv. Its decompressed
bytes, including all 9,600 rows and original line endings, are unchanged.
Every other retained member has exactly the same bytes as the original ZIP.
No new patient-level demographic table or binned age table was added.
Age is not a predictor, stratifier, adjustment variable, endpoint or scoring
input in the included analysis. Event and follow-up times remain unchanged.

CURRENT VERSUS HISTORICAL HASHES
PRIVACY_RELEASE_MANIFEST_20261029.json is the current package inventory.
It records each current payload, the 12 omitted files and the one expanded
text file, with source and current hashes. It excludes only itself from its
file hash inventory. Verify a fresh extraction before running other scripts:
  python release_compatibility_20261029.py

The following unchanged records are retained ONLY as historical provenance:
  README_Scientific_Reports.txt (the earlier 64-file preservation statement)
  SR_PACKAGE_MANIFEST_SHA256.json
  bmc_revision/MANIFEST_SHA256.json
  bmc_revision/PACKAGE_NOTES.json
  bmc_revision/README_REPRODUCE.txt (earlier contents and .joblib inspection)
  bmc_revision/results/reproducibility_manifest.json
  bmc_revision/public_input_verification.json
  bmc_revision/acquisition_reconstruction_test.json
Their inventories describe their original snapshots, not this privacy release.
Historical filenames and hashes do not redistribute the omitted file contents.
Do not use a historical manifest to judge the completeness of this release.

CHECK RESULTS AND REDRAW FIGURES WITHOUT RAW DATA OR MODEL FITTING
In a fresh extraction, install the numerical and plotting dependencies:
  python -m pip install -r bmc_revision/requirements-lock.txt
  python -m pip install -r bmc_revision/requirements-reporting.txt
  python bmc_revision/editorial_result_review.py
  python bmc_revision/make_scientific_reports_figures.py
The first script recomputes external metrics from the saved scores; it writes
only its local editorial audit. The second redraws six PDF/PNG/TIFF figures
from saved results. Neither needs the five omitted CSVs or seven model files.
Do not equate these computational checks with independent clinical validation.

GZIP COMPATIBILITY
No packaged result-reader or plotting script requires the removed gzip path;
benchmark_revision.py writes that path when models are refitted. The released
plain CSV can be read directly with pandas.read_csv(...,
float_precision='round_trip'). If an external historical workflow expects
the gzip filename, this optional command verifies the release and restores
a local gzip copy whose decompressed content is identical:
  python release_compatibility_20261029.py --restore-permutation-gzip
The new gzip header/container hash differs from the original, by design.
It is a LOCAL compatibility output, not a file to upload to medRxiv.

FULL REPRODUCTION IN A SEPARATE LOCAL WORKING COPY
On Windows use a short extraction path, such as C:/VascBridge. A deeply nested
directory can exceed legacy path-length limits while staging GEO source files.
The original input hashes and scientific scripts remain unchanged. Do not
substitute age-redacted CSVs under the original processed-input filenames:
their hashes would fail the frozen downloader's integrity checks. Instead:
  python bmc_revision/download_public_inputs.py
  python bmc_revision/benchmark_revision.py
  python bmc_revision/audit_results.py
The downloader retrieves official NCBI GEO files using public_input_snapshot.json,
or checks byte-identical cached sources, then reconstructs missing expression
and endpoint metadata files in staging and verifies their canonical hashes.
The combined extension_samples_all.csv is not needed by the benchmark; the
unchanged cohort_extension.py can regenerate it locally if separately needed.
The benchmark recreates result sample manifests and all seven serialized
models when the declared models are refitted. joblib remains a required
software dependency, including for its Parallel routine; only model payloads
were removed from the distribution. Full refitting was NOT rerun for this
privacy correction. Public source data remain subject to their source terms.

The official source metadata and locally reconstructed files may again contain
exact ages. Such local analysis inputs, generated model binaries, __pycache__
and gzipped results MUST NOT be zipped wholesale for a new submission. The
historical build_reproducibility_package.py is preserved for scientific/run
provenance but is NOT the medRxiv release builder. Use the dedicated
preparation/build_privacy_release_20261029.py in the author submission folder,
which builds from a hash-locked backup and an explicit omission list, verifies
all retained bytes, and rejects nontext or age-field payloads. It does not
collect a regenerated working directory. The author preparation directory is
not part of the uploaded archive.

SCOPE OF THIS CORRECTION
The release removes the precise-age copies identified in the returned files
and the additional duplicate manifests found during audit. Screening covers
CSV demographic headers and free text, recursive JSON keys, age/DOB patterns,
UTF-8 text decoding, binary signatures and file extensions. It is not a claim
of formal anonymisation or elimination of every possible reidentification risk.
The original scientific limits in the manuscript and README_REPRODUCE.txt
remain applicable. This package does not establish resubmission or posting.
