## Supplementary Data 1: analysis code, provenance and complete outputs for "Cross study transfer of vascular transcriptomic classifiers in seven public cohorts": README_Scientific_Reports.txt

SUPPLEMENTARY DATA 1: VASCBRIDGE COMPUTATIONAL REPRODUCIBILITY
Prepared for Scientific Reports, 25 October 2026

This archive preserves all 64 files from the frozen original reproducibility
archive byte for byte. It adds the plotting recipe used for the six submission
figures, this entry-point guide and an expanded file hash inventory. No models
were refitted and no results were replaced by this packaging operation.

Start with bmc_revision/README_REPRODUCE.txt for the full analysis, data retrieval,
hardware notes and dependency instructions. Public input files are retrieved
from the official GEO/NCBI URLs by the included downloader; the raw expression
files are not redistributed in this compact archive. Saved sample scores,
results, metadata, analysis amendments and hashes are included.

Commands below run from the directory into which this ZIP is extracted.
Use a separate environment (the frozen analysis used Python 3.11).

1. Install numerical and plotting dependencies:
   python -m pip install -r bmc_revision/requirements-lock.txt
   python -m pip install -r bmc_revision/requirements-reporting.txt

2. Check the numerical summaries using saved outputs (no model fitting):
   python bmc_revision/editorial_result_review.py

3. Recreate the six Scientific Reports figures from saved numerical outputs:
   python bmc_revision/make_scientific_reports_figures.py
   Outputs: bmc_revision/scientific_reports_figures/Fig1..Fig6.pdf, .png, .tif
   TIFF files are RGB, 600 dpi, with LZW compression.
   The original make_revision_figures.py remains archived for provenance; it
   produces the earlier layout and is not the current submission figure recipe.

4. To rerun the full public-input analysis, follow README_REPRODUCE.txt. Full
   reruns overwrite results in the extracted working copy. Preserve this ZIP
   and use a fresh extraction when comparing with the frozen reported run.

Interpretation boundaries:
- Seven cohorts contain 294 deposited observations, not 294 independent patients.
- GSE41571 contains 11 macrophage plaque observations (5 ruptured, 6 stable).
- GSE163154 contains 43 segments (27 IPH, 16 non-IPH) from 24 donors; an unresolved
  donor-to-segment crosswalk limits these results to descriptive analysis.
- The common-gene pipeline is separately source-tuned and changes the gene set,
  rank transformation and fitted model together; it is not a one-factor ablation.
- The fixed-baseline permutation control tests source separation, not transport
  or the full model-selection procedure. Exploratory diagnostics were post hoc.
- Computational consistency checks do not constitute external replication,
  human clinical adjudication or evidence for selecting a vascular intervention.

The manuscript and Supplementary Information provide the scientific narrative
and Supplementary Table S1. This ZIP contains computational materials only.
All additions are listed in SR_PACKAGE_MANIFEST_SHA256.json; the older internal
MANIFEST_SHA256.json applies to the original frozen package, not the additions.
