## Supplementary Data 1: analysis code, provenance and complete outputs for "Cross study transfer of vascular transcriptomic classifiers in seven public cohorts": release_compatibility_20261029.py

```
"""Verify the privacy release, optionally restore its text-only gzip locally.

This packaging helper does not download inputs, train models, or change scores.
Run from a fresh extraction before any scientific scripts write audit outputs.
"""
from pathlib import Path
import argparse
import gzip
import hashlib
import json


ROOT = Path(__file__).resolve().parent
MANIFEST = ROOT / 'PRIVACY_RELEASE_MANIFEST_20261029.json'


def sha(data):
    return hashlib.sha256(data).hexdigest()


def main():
    parser = argparse.ArgumentParser(description=__doc__)
    parser.add_argument('--restore-permutation-gzip', action='store_true',
                        help='Create a local gzip copy for historical paths; do not upload the regenerated working directory')
    args = parser.parse_args()
    manifest = json.loads(MANIFEST.read_text(encoding='utf-8'))
    for name, item in manifest['files'].items():
        path = (ROOT / name).resolve()
        path.relative_to(ROOT)
        data = path.read_bytes()
        if len(data) != item['bytes'] or sha(data) != item['sha256']:
            raise RuntimeError('Release-member hash mismatch: ' + name)
    print('PASS: all %d release payload hashes verified' % len(manifest['files']))
    if args.restore_permutation_gzip:
        path = ROOT / 'bmc_revision/results/permutation_sample_predictions.csv'
        destination = path.with_suffix('.csv.gz')
        original = path.read_bytes()
        if destination.exists():
            if gzip.decompress(destination.read_bytes()) != original:
                raise RuntimeError('Existing gzip differs; no file overwritten')
        else:
            with destination.open('xb') as stream:
                with gzip.GzipFile(filename='', mode='wb', fileobj=stream, mtime=0) as compressed:
                    compressed.write(original)
        assert gzip.decompress(destination.read_bytes()) == original
        print('PASS: local gzip decompresses to byte-identical released CSV. '
              'Its container hash need not match the historical gzip. Do not upload this working directory.')


if __name__ == '__main__':
    main()
```
